# GWAS meta-analysis and gene expression data link reproductive tract development, immune response and cellular proliferation/apoptosis with cervical cancer and clarify overlap with other cervical phenotypes

**DOI:** 10.1101/2021.06.18.21259075

**Authors:** Mariann Koel, Urmo Võsa, Maarja Lepamets, Kristi Läll, Natàlia Pujol-Gualdo, Hannele Laivuori, Susanna Lemmelä, Mark Daly, Estonian Biobank Research Team, FinnGen, Priit Palta, Reedik Mägi, Triin Laisk

**Author notes:** Corresponding author: T. Laisk. These authors contributed equally (Mariann Koel, Urmo Võsa).

## Abstract

Genome-wide association studies (GWAS) have successfully identified associations for cervical cancer, but the underlying mechanisms of cervical biology and pathology remain uncharacterised. Our GWAS meta-analyses fill this gap, as we characterise the genetic architecture of cervical phenotypes, including up to 9,229 cases and 490,304 controls for cervical cancer from diverse ancestries. We prioritise *PAX8/PAX8-AS1*, *LINC00339*, *CDC42*, *CLPTM1L*, *HLA-DRB1*, and *GSDMB* as the most likely candidate genes for cervical cancer signals, providing insights into cervical cancer pathogenesis and supporting the involvement of reproductive tract development, immune response, and cellular proliferation/apoptosis. We construct a GRS that associates with cervical cancer (HR=3.7 for top 5% vs lowest 5%), and with other HPV- and immune-system related diagnoses in a PheWAS analysis. Our results propose valuable leads for further functional studies and the presented GRS offers an additional opportunity for risk stratification together with conventional screening strategies.

## Introduction

The uterine cervix has an important role in female reproductive health, as it separates the lower and upper parts of the genital tract and thus forms a barrier for pathogens ^1^ which can cause infection of the upper genital tract. The latter can lead to serious health problems, including pelvic inflammatory disease, infertility, and chronic pelvic pain ^2^. The cervical epithelium is also the main infection site for human papillomavirus (HPV), one of the most common causes of sexually transmitted diseases, which can, in turn, cause cervical dysplasia and in some cases malignant neoplasm of the cervix. Cervical cancer is one of the most common cancer types in women, with more than 28,000 and 311,000 women dying from the disease in Europe and worldwide every year, respectively ^3^.

Although the development of cervical cancer is initiated by high-risk human papillomavirus (HPV) subtype infection, it also involves the contribution from the genetics of the host, which determine whether the infection is successfully cleared or persists and eventually develops into cervical cancer: Host genetics can also influence the rate at which the tumor progresses. Previous family-based studies have estimated the heritability of cervical cancer to be 13-64% ^4–6^ (array-based heritability estimate 7% ^7^), and recent large genome wide association studies (GWAS) have also increased the number of loci reported for cervical cancer ^7, 8^. However, GWAS findings are merely the first step in mapping the genetic susceptibility and biology, and thus, the underlying carcinogenic mechanisms and molecular changes in cervical cancer are still not entirely understood ^9^, nor has the applicability of genetic risk scores in the context of cervical cancer been fully explored.

At the same time, not much is known about the genetic factors modifying other cervical phenotypes, such as cervical ectropion (a benign condition where the columnar epithelium of the cervical canal is turned outwards and exposed to the vaginal environment ^10^), cervicitis (inflammation of the uterine cervical epithelium, most commonly caused by sexually transmitted pathogens, such as *Chlamydia trachomatis*, *Neisseria gonorrhoea* and *Mycoplasma genitalium* ^11^) and cervical dysplasia (a precancerous condition with varying severity, characterized by abnormal growth of the cervical epithelium), which all relate to the cervix and represent partially overlapping conditions with similar symptoms. Without knowing the genetic determinants of other cervical phenotypes and biology, it is difficult to put the findings from cervical cancer genetic studies into the biological perspective.

Here we use data from Estonian Biobank and the FinnGen study to dissect the genetic architecture of cervical phenotypes in a sample set including cases for cervical ectropion (n=10,162), cervicitis (n=19,285), and cervical dysplasia (n=14,694). We then explore their genetic overlap with cervical cancer by combining all publicly available datasets in the largest multi-ancestry GWAS meta-analysis of cervical cancer to date, with 9,229 cervical cancer cases and 490,304 controls. Leveraging latest computational methods and gene expression data, we refine the association signals for cervical cancer and propose potential causal variants and genes at each locus for functional follow-up. Finally, we constructed a genetic risk score, assessed its risk stratification ability and unravelled the pleiotropic phenomic network associating with cervical cancer genetic risk.

## Methods

### Study design and participants

#### Estonian Biobank

The EstBB is a population-based biobank with genotype data and health information for over 200,000 participants ^12^. Information on International Classification of Disease-10 (ICD10) codes is obtained via regular linking with the Health Insurance Fund and other relevant registries ^13^. The 150K data freeze was used for the genetic association analyses described in this paper (n=92,042 women). All biobank participants have signed a broad informed consent for using their data in research and the study was carried out under ethical approval 1.1-12/624 from the Estonian Committee on Bioethics and Human Research (Estonian Ministry of Social Affairs) and data release N05 from the EstBB.

The following ICD10 codes were used for extracting cases: N86 (Erosion and ectropion of cervix uteri), N72 (Inflammatory disease of cervix uteri), N87 (Dysplasia of cervix uteri), C53/D06 (Cervical cancer). Women who did not have the respective ICD codes were used as controls. The final sample size included for analysis was as follows: cervical ectropion: 9,664 cases (average age at joining the biobank ± SD, 35.7 ± 9.6 years), 82,378 controls (45.1 ± 16.3); cervicitis: 18,192 cases (40.9 ± 11.9), 73,850 controls (44.9 ± 16.8); cervical dysplasia: 10,448 cases (39.6 ± 12.1), 81,594 controls (44.6 ± 16.4); cervical cancer 748 cases (50.3 ± 13.4), 81,870 controls (44.6 ± 16.3) (Supplementary Table 1). The overlap of cases for each phenotype can be seen on Supplementary Figure 1. For follow-up analyses, we further stratified the dysplasia cases by severity, resulting in 4,250 cases with mild (N87.0), 2,616 with moderate (N87.1), and 1,599 with severe dysplasia, not elsewhere classified (N87.2), respectively. If more than one diagnosis code was present for dysplasia/cancer, we selected the most severe for analysis (mild < moderate < severe dysplasia < cervical cancer). To validate the cancer diagnosis in the EstBB, we compared the diagnoses for cases (obtained via linking with the National Health Insurance Fund and from self-reported data) to those available through the Estonian Cancer Registry. Reporting of cancer cases to the Cancer Registry is compulsory to all physicians in Estonia who diagnose or treat cancer. Data is also submitted by forensic pathologists. When comparing the same period (diagnoses up to 2016-12-29), out of 707 individuals with C53/D06 diagnosis from other sources, 69% also had the C53/D06 diagnosis in the Cancer Registry. It should be noted that the most recent linking with Cancer Registry includes data up to the end of 2016, therefore the actual overlap is likely higher, as linking with the National Health Insurance Fund has also been done periodically after this date and, for a subset of the individuals, the diagnosis is not yet reflected in the Cancer Registry Data.

All EstBB participants have been genotyped at the Genotyping Core Lab of the Institute of Genomics, University of Tartu, using Illumina Global Screening Array v1.0 and v2.0. Samples were genotyped and PLINK format files were created using Illumina GenomeStudio v2.0.4. Individuals were excluded from the analysis if their call-rate was < 95% or if sex defined based on heterozygosity of X chromosome did not match sex in phenotype data. Before imputation, variants were filtered by call-rate < 95%, HWE p-value < 1e-4 (autosomal variants only), and minor allele frequency (MAF) < 1%. Human genome b37 was used and all variants were changed to be from TOP strand using GSAMD-24v1-0_20011747_A1-b37.strand.RefAlt.zip files from https://www.well.ox.ac.uk/~wrayner/strand/ webpage. Prephasing was done using Eagle v2.3 software ^14^ (number of conditioning haplotypes Eagle2 uses when phasing each sample was set to: --Kpbwt=20000) and imputation was done using Beagle v.28Sep18.79339 with effective population size n_e_=20,000. Population specific imputation reference of 2297 WGS samples was used ^15^. Association analysis was carried out using SAIGE software implementing a mixed logistic regression model ^16^, using year of birth and 10 PCs as covariates in step I.

#### FinnGen study

The FinnGen study is a public-private partnership bringing together genotyping data from different Finnish Biobanks and electronic health records from Finnish health registries. FinnGen release 5 (R5) data, consisting of 218,792 individuals was used, and the following pre-defined phenotypes of interest defined using ICD10, ICD9 and ICD8 codes were used: cervical ectropion (n_cases_=498, n_controls_=68,969), cervicitis (n_cases_=1,093, n_controls_=111,858), cervical dysplasia (n_cases_=4,246, n_controls_=68,969). Additionally, publicly available GWAS summary statistics for phenotype “Malignant neoplasm of cervix uteri” from freeze R4 (n_cases_=1,313, n_controls_=99,048) was used. Since we had only access to summary level data, we have no information on the descriptive statistics (age range) of the sample. In short, phenotypes were defined as follows: ectropion (N14_EROSECTROPUT, ICD10 N86, ICD9 6220A, ICD8 62191), cervicitis (N14_INFCERVIX, ICD10 N72, ICD9 616, ICD 620), dysplasia (N14_DYSPLACERVUT ICD10 N87, ICD9 6221, ICD8 621), cervical cancer (C3_CERVIX_UTERI, ICD10 C53, ICD9 180, ICD8 180). More detailed information on FinnGen endpoint definitions can be found from https://www.finngen.fi/en/researchers/clinical-endpoints. FinnGen individuals were genotyped with Illumina and Thermo Fisher arrays and imputed to the population-specific SISu v3 imputation reference panel according to the following protocol: <u>dx.doi.org/10.17504/protocols.io.xbgfijw</u>. Genetic association testing was carried out with SAIGE ^16^. FinnGen summary statistics included prefiltered variants (minimum allele count>5, imputation INFO score>0.6) and variant positions were converted to b37 using the binary liftOver tool (https://genome.sph.umich.edu/wiki/LiftOver#Binary_liftOver_tool). For more information on genotype data, disease endpoints and GWAS analyses, please see https://finngen.gitbook.io/documentation/. Patients and control subjects in FinnGen provided informed consent for biobank research, based on the Finnish Biobank Act. Alternatively, separate research cohorts, collected prior the Finnish Biobank Act came into effect (in September 2013) and start of FinnGen (August 2017), were collected based on study-specific consents, and later transferred to the Finnish biobanks after approval by Fimea, the National Supervisory Authority for Welfare and Health. Recruitment protocols followed the biobank protocols approved by Fimea. The Coordinating Ethics Committee of the Hospital District of Helsinki and Uusimaa (HUS) approved the FinnGen study protocol Nr HUS/990/2017.

The FinnGen study is approved by Finnish Institute for Health and Welfare (THL), approval number THL/2031/6.02.00/2017, amendments THL/1101/5.05.00/2017, THL/341/6.02.00/2018, THL/2222/6.02.00/2018, THL/283/6.02.00/2019, THL/1721/5.05.00/2019, Digital and population data service agency VRK43431/2017-3, VRK/6909/2018-3, VRK/4415/2019-3 the Social Insurance Institution (KELA) KELA 58/522/2017, KELA 131/522/2018, KELA 70/522/2019, KELA 98/522/2019, and Statistics Finland TK-53-1041-17.

The Biobank Access Decisions for FinnGen samples and data utilized in FinnGen Data Freeze 5 include: THL Biobank BB2017_55, BB2017_111, BB2018_19, BB_2018_34, BB_2018_67, BB2018_71, BB2019_7, BB2019_8, BB2019_26, Finnish Red Cross Blood Service Biobank 7.12.2017, Helsinki Biobank HUS/359/2017, Auria Biobank AB17-5154, Biobank Borealis of Northern Finland_2017_1013, Biobank of Eastern Finland 1186/2018, Finnish Clinical Biobank Tampere MH0004, Central Finland Biobank 1-2017, and Terveystalo Biobank STB 2018001.

#### Publicly available datasets

For cervical cancer meta-analysis we additionally used publicly available datasets from Rashkin et al 2020 ^7^ (downloaded from https://github.com/Wittelab/pancancer_pleiotropy, including 5,998 cases and 189,855 controls from the UK Biobank, and 565 cases and 29,801 controls from Kaiser Permanente cohort) and summary statistics from Biobank Japan ^17^ (downloaded from http://jenger.riken.jp/en/result), including 605 cases and 89,731 controls. The summary statistics include variant-level association statistics (effect estimate and standard error (SE), association p-value etc.). For the data from Rashkin et al study, we first converted OR-s to betas (beta=log(OR)), then derived z-scores from reported p-values (using the ‘qnorm’ function in R) and calculated SE-s (SE=beta/z-score).

#### GWAS meta-analysis

All European ancestry meta-analyses were conducted using inverse variance weighted fixed-effect meta-analysis method implemented into GWAMA software (v2.2.2) ^18^. For cervical cancer meta-analysis including data from Biobank Japan, we used MR-MEGA, which is a tool for multi-ancestry meta-regression ^19^. Genome-wide significance was set at p < 5 × 10^-8^ in all analyses. We used MTAG v1.0.8 ^20^ (Multi-Trait Analysis of GWAS) to jointly analyse the summary statistics from dysplasia and cervical cancer European ancestry analyses and thus increase the power to detect additional associations.

Variant annotation and follow-up analyses were done using individual trait GWAS summary statistics from European ancestry analyses.

#### Annotation of GWAS signals

We used FUMA v.1.3.6 ^21^ for functional annotation of GWAS results and credible set variants. For functional annotation, the Annotate Variation (ANNOVAR) ^22^, CADD (a continuous score showing how deleterious the SNP is to protein structure/function; scores >12.37 indicate potential pathogenicity)^23^ and RegulomeDB ^24^ scores (ranging from 1 to 7, where lower score indicates greater evidence for having regulatory function), and 15 chromatin states from the Roadmap Epigenomics Project ^25^ were used. FUMA also performs lookups in the GWAS Catalog (e96_r2019-09-24), the results of which are shown in Supplementary Table 7 and Supplementary Figure 11.

#### Look-up of variants previously associated with cervical cancer

We used our European ancestry only and multi-ancestry cervical cancer meta-analysis summary statistics to conduct a look-up of variants previously reported in association with cervical carcinoma. For this, we extracted variants associated with the EFO term EFO_0001061(cervical carcinoma) from the GWAS Catalog. The results of this look-up can be seen in the Supplementary Table 1.

#### Gene-based tests

We used MAGMA (v1.08) implemented in FUMA with default settings to conduct gene-based genome-wide association testing. According to the number of tested protein-coding genes, the genome-wide significance level was set at 0.05/19,913=2.7 × 10^-6^.

#### HLA analysis

For cervical dysplasia meta-analysis, we carried out HLA imputation of the EstBB genotype data with the SNP2HLA v1.0.3 tool ^26^. As an imputation reference, we used a merged reference of EstBB WGS ^15^ and Type 1 Diabetes Genetics Consortium samples ^26^. We tested for association between the alleles and cervical dysplasia in the EstBB using SAIGE with the LOCO option. We used imputed data on alleles (two- and four-digit) in the MHC class I genes (*HLA-A*, *HLA-B*, *HLA-C*) and classical MHC class II genes (*HLA- DRB1*, *HLA-DQA1*, *HLA-DQB1*, *HLA-DPA1*, *HLA-DPB1*) for 10,446 cases and 81,586 controls in the EstBB, who had the corresponding data available.

#### Colocalisation and fine-mapping analyses

We used HyPrColoc (v1.0.0) ^27^, a fast and efficient colocalisation method for identifying the overlap between our GWAS meta-analysis signals and *cis*-QTL signals from different tissues and cell types (expression QTLs, transcript QTLs, exon QTLs and exon usage QTLs available in the eQTL Catalogue ^28^). We lifted the GWAS summary statistics over to hg38 build to match the eQTL Catalogue using binary liftOver tool (https://genome.sph.umich.edu/wiki/LiftOver#Binary_liftOver_tool). For each genome-wide significant (p<5 × 10^-8^) GWAS locus we extracted the +/-500kb of its top hit from QTL datasets and ran the colocalization analysis against eQTL Catalogue traits. For each eQTL Catalogue dataset we included all the QTL features which shared at least 80% of tested variants with the variants present in our GWAS region. We used the default settings for HyPrColoc analyses and did not specify any sample overlap argument, because HyPrColoc paper ^27^ demonstrates that assuming trait independence gives reasonable results. HyPrColoc outputs the following results a) a cluster of putatively colocalised traits (here our GWAS region of interest and *cis*-QTL signal for any nearby feature for given QTL dataset); b) the posterior probability that genetic association signals for those traits are colocalising (we considered two or more signals to colocalize if the posterior probability for a shared causal variant (PP4) was 0.8 or higher. All results with a PP4 > 0.8 can be found in Supplementary Table 4); c) the ‘regional association’ probability (a large regional association probability indicates that one or more SNPs in the region have shared association across evaluated traits); d) a candidate causal variant explaining the shared association; and e) the proportion of the posterior probability explained by this variant (which also represents the HyPrColoc multi-trait fine-mapping probability). For every colocalisation event, we also calculated 95% credible set (CS) for multi-trait fine-mapping results. To do so, we ranked all variants decreasingly based on their posterior probability and extracted top *n* variants with cumulative posterior probability of ≥0.95.

Since cervical samples were not present in analysed gene expression datasets, we prioritised colocalisation signals from tissues that cluster together with vagina/uterus in GTEx V8 data, either based on cell-type-composition or gene expression (Supplementary figures S41 and S48 of ^29^. These tissues include vagina, uterus, esophagus mucosa and gastro-esophageal junction, sigmoid colon, skin, salivary gland, and tibial nerve. Of these ‘proxy’ tissues, esophageal mucosa (stratified squamous epithelium) and gastro-esophageal junction (transition zone between stratified and columnar epithelium) tissues are histologically most similar to the cervix.

We used FUMA ^21^ to annotate credible set variants with chromatin 15-state marks in HeLa-S3 Cervical Carcinoma cell line (E117) and in available ‘proxy ’ tissues (E106 - sigmoid colon; E079 - esophagus; E055-E061,E126, E127 - skin) from the Roadmap Epigenomics Project ^25^.

#### Genetic correlations

We used the LD Score regression method ^30^ implemented in LD Hub ^31^(http://ldsc.broadinstitute.org) for testing genetic correlations between cervical cancer and traits spanning reproductive, aging, autoimmune, cancer, and smoking behaviour categories (33 traits in total), using the cervical cancer European-ancestry only GWAS meta-analysis summary statistics and data available within the LD Hub resource. After filtering the input to HapMap3 SNPs, removing SNPs within the MHC region, and merging with the built-in reference panel LD Scores (1000 Genomes EUR ancestry) ^31^, ∼1.1M variants remained for analysis. False discovery rate (FDR) correction (calculated using the p.adjust function in R) was used to account for multiple testing. Results of the analysis are presented in Supplementary Table 9.

LDSC estimated observed scale heritability (0.0059 (se=0.0013)) for cervical cancer was converted to liability scale using the formula h_2liability_ = h_2observed_ × K^2^ × (1 - K)^2^ / P / (1-P) / zv^2^, where K is the population prevalence (here equal to sample prevalence) and P is the proportion of cases in the study (European ancestry analysis, 2.1%). This resulted in a liability scale heritability estimate of 4.75% for non-HLA common variant heritability.

#### Genetic risk score analysis

We constructed a GRS for cervical cancer based on the summary statistics of the meta-analysis including the summary statistics from Rashkin et al and FinnGen, with 7,876 cases and 318,704 controls of European ancestry, leaving out EstBB 200K dataset as an independent target dataset (1,094 cervical cancer cases, 131,314 female controls).

We computed and evaluated ten versions of GRS for each individual in the EstBB 200K dataset (132,408 women, 70,502 men) implementing LDPred ^32^, which uses a linkage-disequilibrium SNP-reweighting approach. The following fractions of causal variants were used: 1, 0.3, 0.1, 0.03, 0.01, 0.003, 0.001, 0.0003, 0.0001.LD data from Estonian WGS dataset (n=2297) was used for reference. STEROID (v0.1.1) was used for calculating PRS for all EstBB participants (https://genomics.ut.ee/en/tools/steroid).

First, we divided the target EstBB dataset into a discovery (prevalent cases) and validation (incident cases) dataset. The discovery dataset included 859 prevalent cases and 3,436 controls (4 controls per case). Since controls were defined as women who did not develop cervical cancer during follow-up, they tended to be younger than prevalent cases. We used the discovery set to select the best predicting PRS version using a logistic regression model adjusted for age, age squared and smoking status (coded as “Never”, “Former”, “Current”). We used smoking status as a covariate, as it is a known risk factor for cervical cancer and was easily available for all included biobank participants. The GRS that had the smallest p-value in discovery set analysis, which contained 2,894,555 variants, was selected for further analyses.

The validation set included 235 incident cases and 127,878 controls, and in this set we tested the predictive ability of GRS (Supplementary Figure 2). We standardised the best PRS version and also categorized it into different percentiles (<5%, 5%-15%, 15%-25%, 25%-50%, 50%-75%, 75%-85%, 85%-95%, >95%). Cox proportional hazard models were used to estimate the Hazard Ratios (HR) corresponding to one standard deviation (SD) of the continuous GRS for the validation dataset. Harrell’s C-statistic was used to characterize the discriminative ability of each GRS. Cumulative incidence estimates were computed using Kaplan-Meier method and to account for competing events (mortality), we used the “cmprsk” R library. While comparing different GRS groups with each other, age was used as a timescale to properly account for left-truncation in the data.

To explore how much of the GRS predictive power comes from the HLA region, we separated the GRS into HLA and non-HLA fractions. We took the best performing GRS and extracted the markers and LDpred weights in the HLA region (chr6:28,477,797- 33,448,354, number of variants = 9,764) and calculated separate scores for the HLA region using STEROID. We then subtracted the HLA score from the overall score, resulting in the nonHLA score.

We also performed a pheWAS analysis with the best-performing GRS, where we tested the association between the GRS and all ICD10 diagnosis codes in EstBB 200K data (excluding relatives using a pihat cut-off value 0.2) in a logistic regression framework, adjusting for sex, age, and ten PCs. Separate analyses stratified by sex were also performed. Bonferroni correction was applied to select statistically significant associations (number of tested ICD maincodes - 2001, corrected p value threshold 0.05/2001=2.5 × 10^-5^. We repeated the overall pheWAS analysis with the HLA and nonHLA scores to clarify which fraction of the score drove the associations. Results were visualized using the PheWas library (https://github.com/PheWAS/PheWAS). All analyses were carried out in R 3.6.1 or R 4.1.1.

## Results

First, to determine the genetic factors associated with cervical phenotypes, we conducted GWAS using 92,042 female individuals from the EstBB for cervical ectropion, cervicitis, and dysplasia. Next, the results of these analyses were further meta-analysed together with the corresponding summary statistics from the FinnGen study (R5 release data with - depending on the phenotype - up to 112,951 Finnish female individuals). The resulting meta-analysis included 10,162 women with cervical ectropion, 19,285 with cervicitis, 14,694 with cervical dysplasia and up to 193,452 female controls of European ancestry.

### GWAS meta-analyses for cervical ectropion, cervicitis, and dysplasia

We identified one genome-wide significant (p < 5 × 10^-8^) locus for both cervical ectropion and cervicitis (Supplementary Figure 3), and five signals for cervical dysplasia (**Table 1;** Supplementary Figure 4) (number of analysed markers in meta-analysis up to 11,043,697). All the reported genetic variants show at least nominal significance in both analysed cohorts (**Table 1**).

**Table 1.** GWAS meta-analyses results. Genetic variants associated with cervical ectropion, cervicitis, and dysplasia.

| Phenotype | chr:pos<br>(b37) | Variant<br>(EA) |  | Meta-analysis |
| --- | --- | --- | --- | --- |
| Ectropion | 2:113984033 | rs3748916<br>(A) | P-value<br>OR (95% CI)<br>EAF | $5.1 \times 10^{-16}$<br>1.14 (1.20-1.18)<br>0.41 |
| Cervicitis | 2:113975110 | rs1049137<br>(G) | P-value<br>OR (95% CI)<br>EAF | $3.9 \times 10^{-10}$<br>0.92 (0.91-0.94)<br>0.25 |
| Dysplasia | 2:113975110 | rs1049137<br>(G) | P-value<br>OR (95% CI)<br>EAF | $6.4 \times 10^{-9}$<br>0.92 (0.91-0.94)<br>0.26 |
| | 2:159629994 | rs12611652<br>(A) | P-value<br>OR (95% CI)<br>EAF | $3.2 \times 10^{-9}$<br>0.92 (0.91-0.94)<br>0.54 |
| | 5:1311693 | rs6866294<br>(C) | P-value<br>OR (95% CI)<br>EAF | $2.1 \times 10^{-9}$<br>1.08 (1.06-1.10)<br>0.57 |
| | 6:31322047 | rs1053726<br>(G) | P-value<br>OR (95% CI)<br>EAF | $9.1 \times 10^{-9}$<br>0.91 (0.88 - 0.95)<br>0.19 |
| | 6:32611759 | rs36214159<br>(G) | P-value<br>OR (95% CI)<br>EAF | $1.6 \times 10^{-22}$<br>0.79 (0.76-0.83)<br>0.09 |

Notably, all three analysed phenotypes showed significant association with a locus on chromosome 2 near *PAX8* gene, a transcription factor known to be relevant for genital tract development and its antisense RNA *PAX8-AS1*.

Furthermore, we observed additional four genome-wide significant signals for cervical dysplasia - two in the HLA region on chromosome 6 (rs1053726, p=9.1 × 10^-9^, rs36214159, p=1.6 × 10^-22^), one on chromosome 2 (rs112611652, p=3.2 × 10^-9^) near *DAPL1* and one on chromosome 5 (rs6866294, p=2.1 × 10^-9^), downstream *CLPTM1L*.

### GWAS meta-analysis for cervical cancer

To determine the genetic overlap between cervical phenotypes and cervical cancer, we conducted another GWAS analysis with the EstBB data for cervical cancer (n_cases_=748) and combined the results with publicly available GWAS summary statistics resulting in the largest GWAS meta-analysis for cervical cancer to date. We used data from FinnGen release R4 (n_cases_=1,313; https://r4.finngen.fi/pheno/C3_CERVIX_UTERI), Rashkin et al 2020 ^7^ (n_cases_=6,563) and Biobank Japan (n_cases_=605; http://jenger.riken.jp:8080/pheno/Cervical_cancer), resulting in a total of 8,624 cases and 400,573 controls for the European ancestry meta-analysis, and totalling 9,229 cervical cancer cases and 490,304 controls in the multi-ancestry meta-analysis.

As a result, we identified five loci associated with cervical cancer (**Table 2;** Supplementary Figure 5): 1p36.12 (rs2268177, p= 3.08 × 10^-8^), 2q13 (rs4849177, p=9.36 × 10^-15^), 5p15.33 (rs27069, p=1.31 × 10^-14^), 17q12 (rs12603332, p=1.18 × 10^-9^), and in the HLA region on 6p21.32 (multi-ancestry meta-analysis: rs35508382, p=1.04 × 10^-39^; European ancestry analysis: rs28718232, p=2.55 × 10^-44^), with similar effect estimates in European ancestry and Biobank Japan datasets (**Table 2**). We then proceeded to define the most likely causal SNPs and the most likely causal gene at each associated locus (except the HLA region, for which we conducted separate signal finemapping, see below) using the European ancestry meta-analysis results. We considered the following criteria when selecting the most likely candidate genes (**Figure 1**) - a) whether the lead signal is in LD with a coding variant in any of the nearby genes, b) which is the closest gene to GWAS lead variant in each locus, and c) is there significant (posterior probability >0.8) colocalisation in relevant tissues (tissues similar to female reproductive tract tissues based on cellular composition and gene expression). For credible set variants, we highlighted those that have a larger regulatory potential based on the HeLa cell line data (Supplementary Figure 6, Supplementary Table 6).

**Figure 1.**
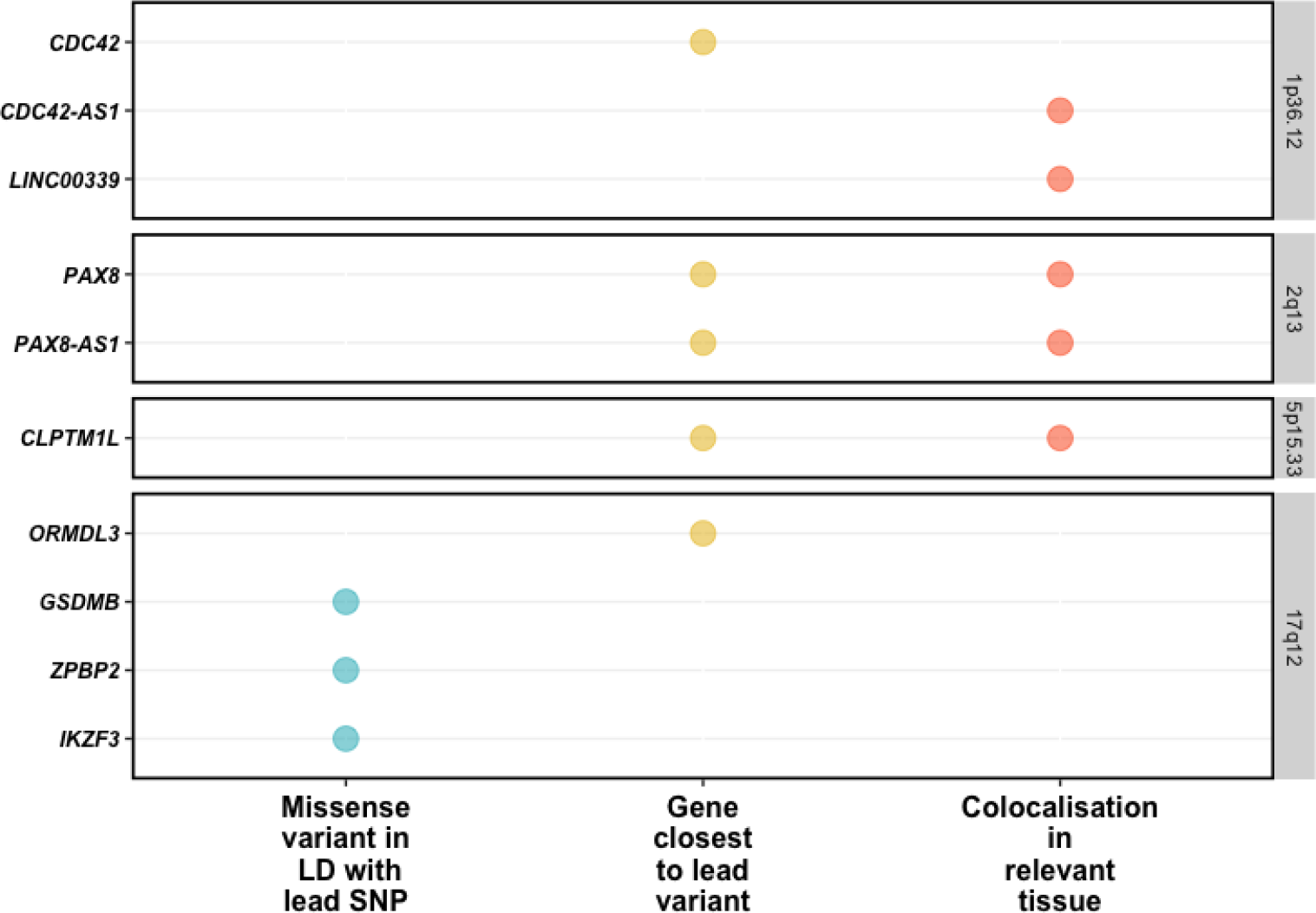
Visualised evidence for cervical cancer GWAS meta-analysis candidate gene mapping (showing only genes with at least one level of evidence). We considered the following criteria when selecting the most likely candidate genes - a) whether the lead signal is in LD (r^2^ >0.6) with a coding variant in any of the nearby genes, b) which is the closest gene to GWAS lead variant in each locus, and c) is there significant (posterior probability >0.8) colocalisation in relevant tissues (tissues similar to female reproductive tract tissues based on cellular composition and gene expression.

**Table 2.**
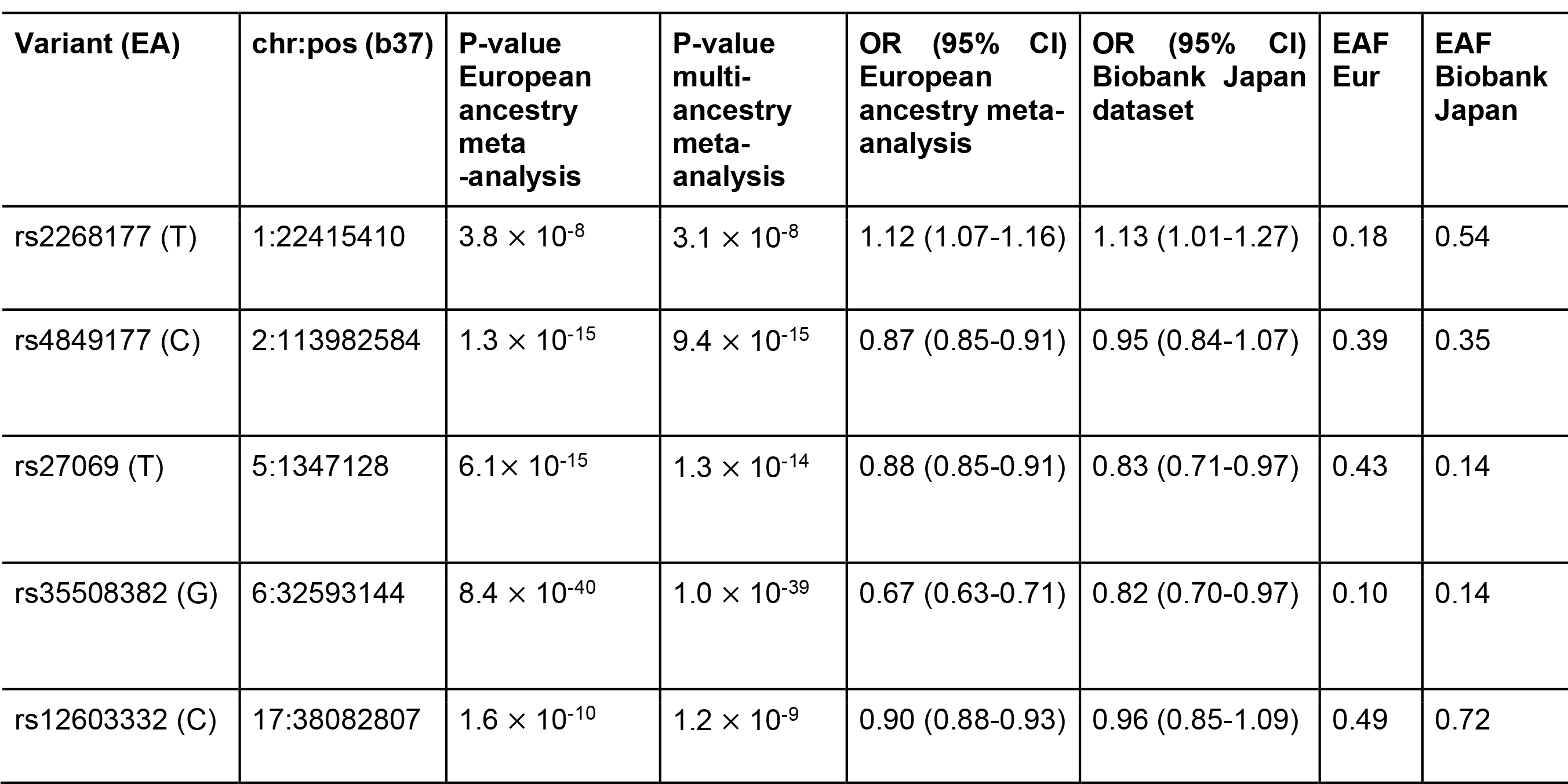
Genetic markers associated with cervical cancer European ancestry and multi-ancestry meta-analyses

The lead variant on 1p36.12 (rs2268177) is in the intron of *CDC42*, downstream *WNT4*. Colocalisation analysis showed that cervical cancer GWAS association signal colocalises with *CDC42*, *CDC42-AS1* and *LINC00339* expression/transcription events in several tissues and cell types (Supplementary Table 3) and corresponding credible sets included 40 variants (Supplementary Table 4). *CDC42-AS1* and *LINC00339* were also prioritised based on the colocalisation signal in trait-relevant tissue (**Figure 1**), with high colocalisation probability (PP4=0.94) between the GWAS signal and *CDC42-AS1* gene expression in esophagus mucosa, and between the GWAS signal and *LINC00339* transcripts ENST00000635675 and ENST00000434233 in GTEx skin dataset. In both colocalisations, rs2473290 (in the intron of *CDC42-AS1*) explains most of the shared association (posterior inclusion probability 0.95-0.99). Of the other credible set variants, rs3768579 and rs3754496 are located in transcription start site (TSS) flanking regions of *LINC00339* and *CDC42* in HeLa cells, while rs72665317 and rs10917128 overlap with enhancer marks (Supplementary Table 6, Supplementary Figure 6). *LINC00339* has a known role in promoting the proliferation of several cancers ^33–35^, while there is also evidence to link *CDC42* expression with cervical cancer invasion and migration ^36^. The region has been previously associated with uterine fibroids, endometriosis, endometrial cancer ^37^, epithelial ovarian cancer, gestational age, and bone mineral density (Supplementary Table 7).

As with other cervical phenotypes, we observed a significant association on chromosome 2, where the lead genetic marker (rs4849177) is in an intronic region of *PAX8*. The GWAS signal colocalises with the expression of *PAX8* and its potential regulator, *PAX8-AS1*, in several tissues and cell types, and the credible set included 29 variants. Of the credible set variants, rs1015753 overlaps with a TSS flanking region in HeLa cells, while another six variants overlap with regulatory enhancer elements (Supplementary Table 6). Colocalisation signals for *PAX8* and *PAX8-AS1* were also observed in several relevant tissues, including vagina (Supplementary Table 3), where the credible set included 13 variants (Supplementary Table 4), two of them overlapping with enhancer elements.

We compared the signal in the 2q13 locus across the analysed cervical phenotypes (Supplementary Figures 7 and 8; Supplementary Table 5) and found that the lead signals for ectropion (rs3748916) and cervicitis/dysplasia (rs1049137) are not in high linkage disequilibrium (r2=0.27, 1000G p3v5 EUR), indicating independent or partly independent signals in the same region. The cervical cancer lead signal was moderately correlated (r2=0.45-0.53, EUR) with cervicitis/dysplasia and ectropion signals, respectively. This is supported by the fact that although the sets of most likely causal variants mostly overlapped for cervicitis, dysplasia, and cancer, the credible set variants seem to be different for ectropion.

The signal on chromosome 5 (lead variant rs27069) locates upstream *CLPTM1L* and overlaps with the TSS in HeLa cells (Supplementary Figure 6). Numerous colocalisations with different *CLPTM1L* QTL events were observed, including in skin and gastroesophageal junction datasets (Supplementary Table 3). *CLPTM1L* is a membrane protein and its overexpression in cisplatin-sensitive cells causes apoptosis. Polymorphisms in this region have been reported to increase susceptibility to cancer, including lung, pancreatic, and breast cancers (Supplementary Table 7). Variants in the credible set overlap with active TSS, as well as with several enhancer and ZNF repeat marks in the *CLPTM1L* gene (Supplementary Figure 6).

On chromosome 17, the lead signal (rs12603332) is in high LD (r2>0.8) a splice acceptor variant (rs11078928) in *GSDMB*. GSDMB belongs to the family of gasdermin-domain containing proteins. Members of this family regulate apoptosis in epithelial cells and are linked to cancer ^38^. GSDMB has also been linked with invasion and metastasis in breast cancer cells ^39^ and in cervical cancer ^40^. Specifically, the splice variant rs11078928 deletes exon 6 which encodes 13 amino acids in the critical N-terminus, and therefore abolishes the pyroptotic activity (pyroptosis is a type of cell death) of the GSDMB protein^41^. This region has been previously associated with asthma, inflammatory bowel disease, ulcerative colitis, Crohn’s disease, multiple sclerosis, primary biliary cholangitis, rheumatoid arthritis and other disorders with an immune etiology, but also with cervical cancer ^42^.

Given the similarity in signals identified for cervical dysplasia and cervical cancer (Table 1 and 2), we jointly analysed the GWAS results for dysplasia and cancer and identified an additional signal on chromosome 19 (rs425787, p=3.5 ×10^-8^, Supplementary Figure 9) that remained below the significance threshold in the cervical cancer analysis alone (p=2.1×10^-7^). Since this locus was not significant in the cervical cancer meta-analysis, it was not included in the colocalisation and fine-mapping analyses. This association signal overlaps with enhancer histone marks in HeLa cervical carcinoma cell line (Supplementary Figure 9) and is in the 3’ region of *CD70*. CD70 is a cytokine with an important role in T-cell immunity during antiviral response, and its high expression has been associated with a favourable outcome in cervical cancer patients ^43^.

### Dysplasia signals stratified by dysplasia severity and in cervical cancer

We stratified the dysplasia phenotype to evaluate the meta-analysis effect sizes (odds ratios) in relation to pathology severity. Figure 2 shows the effect estimates in dysplasia subphenotypes and in cervical cancer meta-analysis from European ancestry. In general, odds ratios correlated with degree of pathology, although there was an overlap in confidence intervals (Figure 2). An interesting exception seems to be rs12611652 near *DAPL1*, which is associated with different cervical dysplasia subphenotypes, but not with cervical cancer. *DAPL1* is expressed in epithelium and may play a role in the early stages of epithelial differentiation or in apoptosis and is a suppressor of cell proliferation in retinal pigment epithelium ^44^. The GWAS signal colocalises with *PKP4* expression in gastro- esophageal junction tissue, with three SNPs in the credible set (Supplementary Table 8). PKP4 regulates junctional plaque organisation, cadherin function, and cell adhesion.

**Figure 2.**
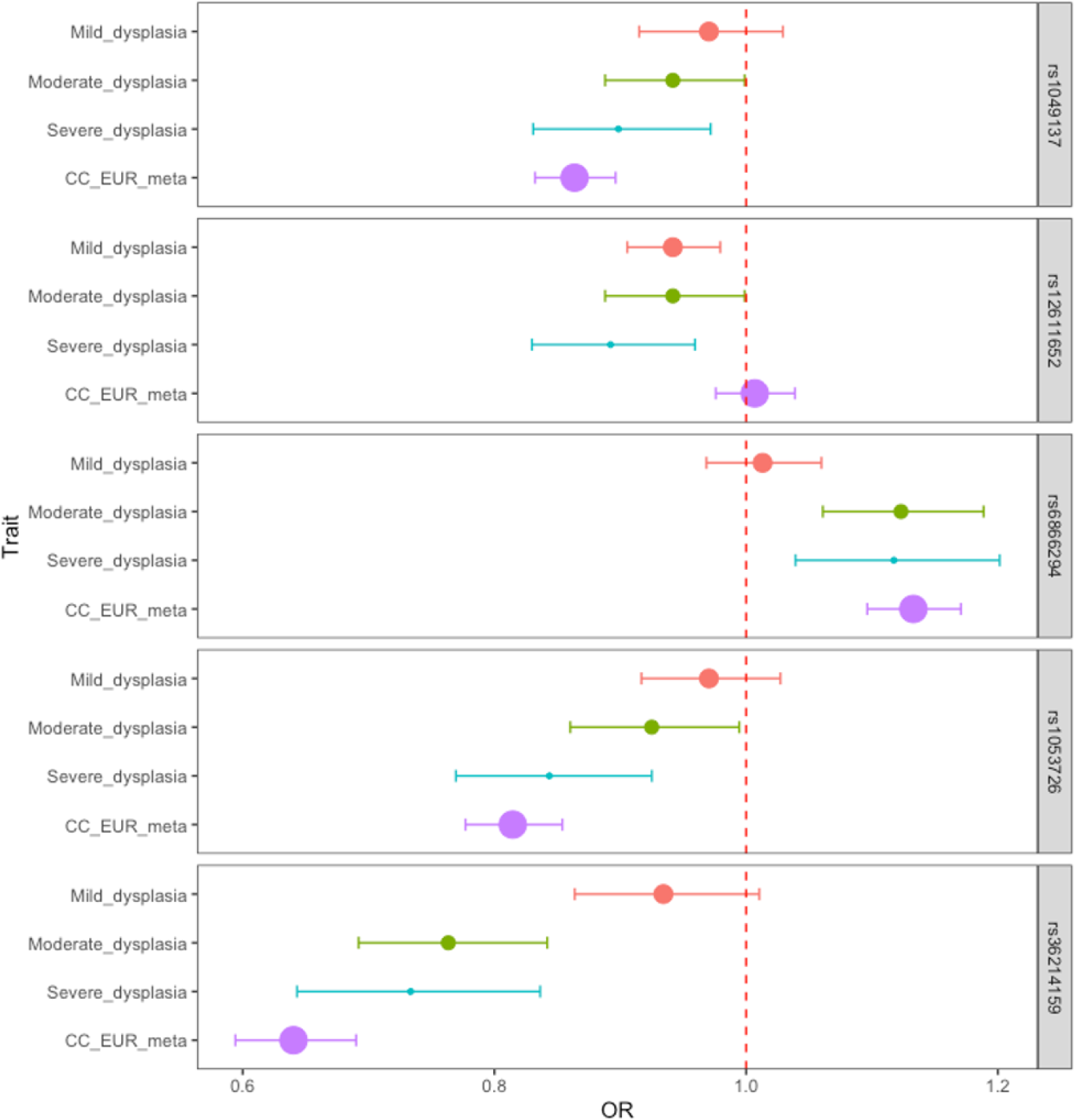
Dysplasia lead signals in different dysplasia stages and cervical cancer in European ancestry analyses. Data are presented as odds ratios (dot) and 95% confidence intervals (error bars) originating from GWAS analysis. The size of the dot is proportional to the effective sample size (calculated as 4/((1/N_cases)+(1/N_controls)). The red dashed line represents the line of no effect.

**Figure 3.**
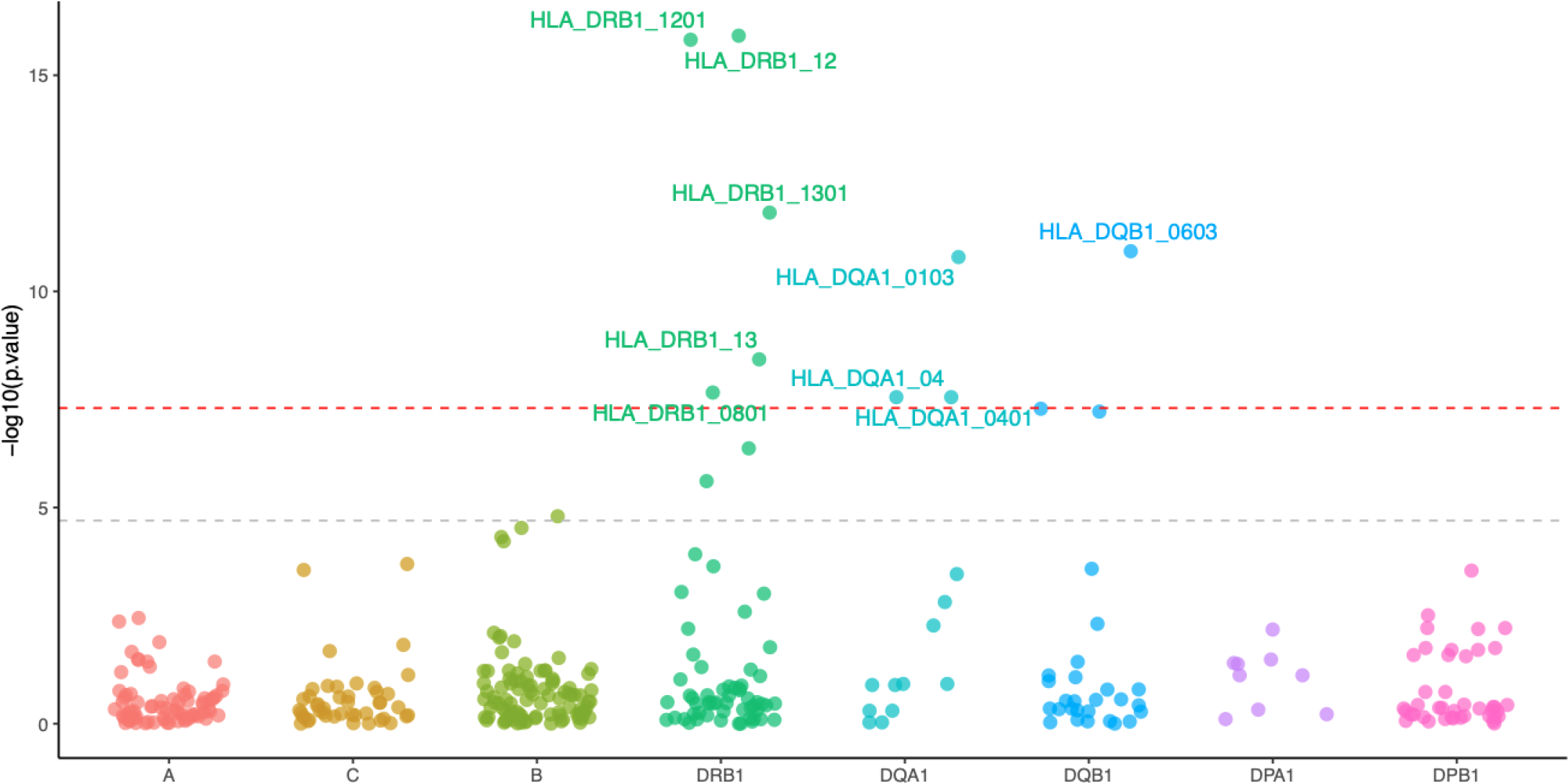
HLA alleles associated with cervical dysplasia. The y-axis shows the -log10 P-values from analysis of 10,446 cases and 81,586 controls in the EstBB using SAIGE. The red dashed line represents the genome-wide significance threshold (p<5 × 10^-8^), while the grey line represents the p-value threshold adjusted for the number of tested alleles (p<2.0 × 10^-5^).

All cervical cancer lead signals were at least nominally significant (p<0.05) in cervical dysplasia analysis, rs4849177 and rs35508382 were also genome-wide significant (Supplementary Table 2), confirming the overlap of genetic risk factors for cervical dysplasia and cancer.

### Gene-based testing in cervical cancer

The results from gene-based testing largely mirror the results of single variant analysis. Apart from numerous genes on chromosome 6, *CLPTM1L*, *PAX8*, *PSD4*, *GSDMB*, *ORMDL3*, *ZPBP2*, *CD70*, and *SKAP1* passed the significance level threshold (p<2.5 × 10^-6^, Supplementary Table 10), with the association with *SKAP1* being a novel finding compared to single variant analysis. SKAP1 is a T-cell adaptor protein with a critical role in coupling T-cell antigen receptor stimulation to the activation of integrins. *The SKAP1* locus has been previously associated with ovarian cancer ^45^.

### Look-up of variants previously associated with cervical cancer

Of the 170 variants with an rs-number extracted from GWAS catalog as (potentially) associated (p-value < 9 × 10^-6^) with cervical cancer, 55 were present in all four of the cohorts included in the multi-ancestry meta-analysis. Of these, 34 had a p-value <9.1 × 10^-4^ (Supplementary Table 11), which is the Bonferroni corrected threshold of significance (0.05/55). In the European ancestry analysis, 64/170 variants were present and 19 passed the Bonferroni corrected threshold of significance (0.05/64=7.8 × 10^-4^), including variants in/near *PAX8*, *MUC21/MUC22*, the *HLA* gene cluster, and *GSDMB*.

### HLA fine-mapping

Since both cervical dysplasia and cervical cancer show an association signal in the HLA region, we used the larger cervical dysplasia dataset in EstBB to further map the cervical dysplasia association signal in the HLA region. *HLA-DRB1*\*1201 (p=1.2 × 10^-16^, OR=0.74 (0.68-0.79)), *HLA-DRB1*\*1301 (p=1.5 × 10^-11^, OR=0.82 (0.78-0.87)), *HLA-DQB1*\*0603 (p=1.2 × 10^-11^, OR=0.83 (0.79-0.88)), *HLA-DQA1*\*0103 (p=1.6 × 10^-11^, OR=0.83 (0.79-0.88)), *HLA-DRB1*\*0801 (p=2.2 × 10^-8^, OR=1.20 (1.12-1.27)) and *HLA-DQA1*\*0401 (p=2.8 × 10^-8^, OR=1.19 (1.12-1.27)) alleles passed the genome-wide significance threshold. These results are in line with previous studies in cervical cancer - *HLA*-*DRB1*\*1301 and *DQB1*\*0603 alleles are associated with decreased risk ^46–49^, and more broadly, the *HLA*-*DRB1*\*1301–*HLA*-*DQA1*\*0103–*HLA*-*DQB1*\*0603 haplotype has been shown to protect against cervical cancer ^50^. *HLA*-*DRB1*\*0801 and *HLA*-*DQA1*\*0401 are in strong LD with *HLA*-*DQB1*\*0402 (p=5.2x10^-8^) and have been associated with autoimmune disease, including type 1 diabetes and systemic lupus erythematosus ^51^.

### Genetic correlations

We evaluated pairwise genetic correlations (r_g_) between cervical cancer and 33 selected traits from LD Hub. We found two significant (FDR<0.05) genetic correlations - age at first birth (r_g_=-0.37, se=0.08) and former vs current smoking status (r_g_=-0.45, se=0.14). Several other traits reflective of smoking behavior (incl. lung cancer) were also nominally significant (Supplementary Table 9).

### Genetic risk score for cervical cancer

Evaluating a total of ten risk score profiles, we found the best performing score had an OR=1.45 (95% CI 1.32 - 1.59, p= 1.68 × 10^-14^) for discriminating between case/control status in the discovery stage (Supplementary Figure 11). Cervical cancer prevalence in EstBB according to genetic risk categories can be seen on Figure 4.

**Figure 4.**
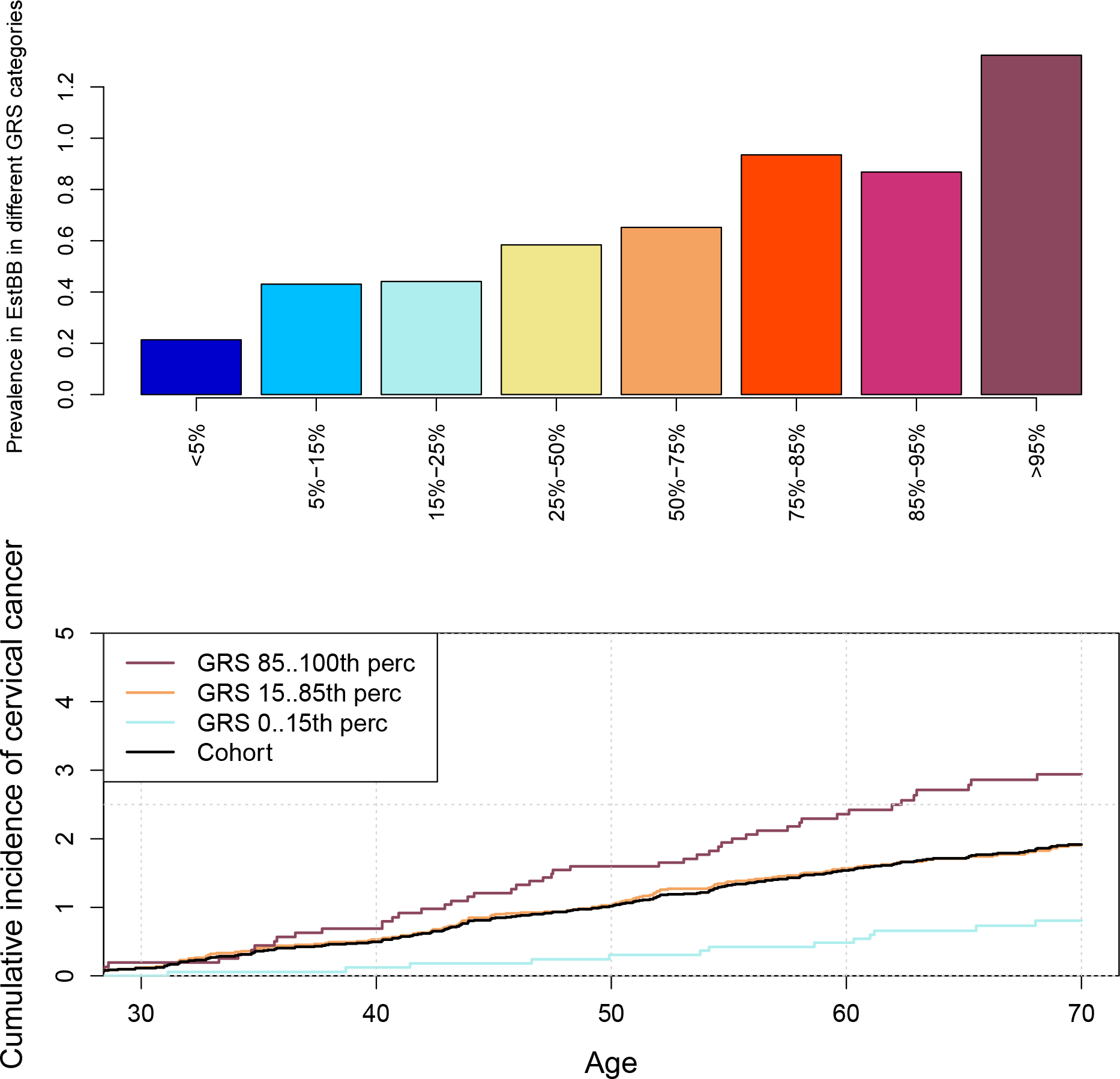
Prevalence of cervical cancer in different genetic risk categories (top) and cumulative incidence in % according to genetic risk (bottom) in EstBB. Cumulative incidence accounting for competing risk in three GRS categories, in women up to 70 years. Black line represents the cohort average.

**Figure 5.**
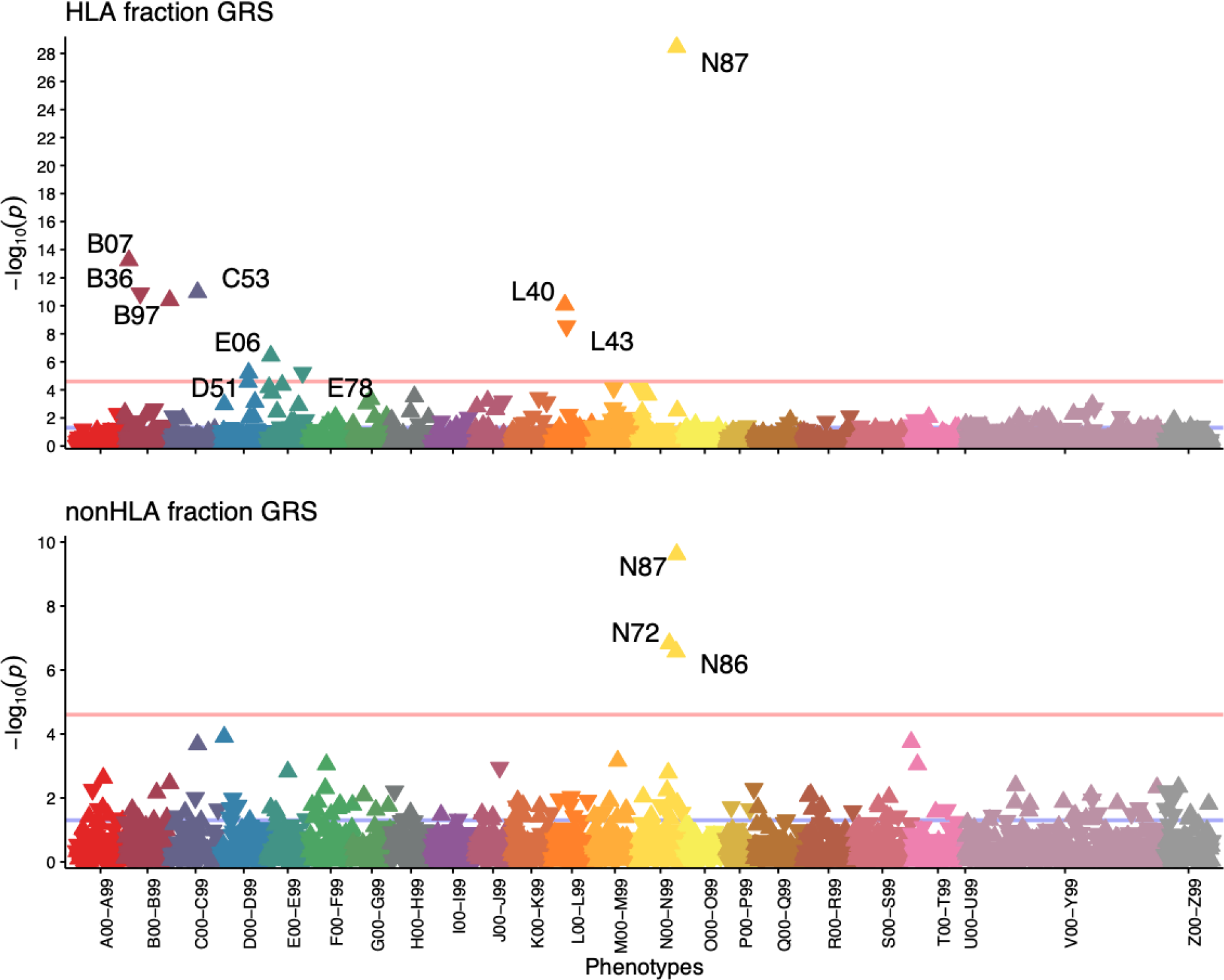
pheWAS results for association with cervical cancer risk score HLA (top) and nonHLA (bottom) fractions. Each triangle in the plot represents one ICD10 main code and the direction of the triangle represents direction of effect—upward-pointing triangles show increased probability of a diagnosis code in individuals with higher GRS. Pink line—Bonferroni-corrected significance level (p=2.5 × 10^−5^).

The HLA fraction of the score had an OR=1.35 (1.23-1.48) and the nonHLA fraction OR=1.25 (1.14-1.37), indicating that the majority of the predictive power comes from the *HLA* region, with marginal contribution from the rest of the genome.

We then evaluated the performance of this GRS in the validation set, consisting of incident cervical cancer cases (n=235) and controls (n=127,878). In the validation set, the risk increased 1.33-fold per 1SD increase of risk score and the continuous distribution of GRS showed a C-statistic of 0.61 (SD=0.02). Then, we divided the GRS into following categories: <5%, 5%-15%, 15%-25%, 25%-50%, 50%-75%, 75%-85%, 85%-95%, >95%, and <15%, 15-85%, >85%. Cumulative incidence of cervical cancer according to genetic risk category while accounting for competing events (death) can be seen on Figure 4. For women in the top 5% risk group, the cervical cancer rate was 3.7 times as great as that for the individuals in the lowest 5%.

In order to interpret our findings and assess the overlap with other phenotypes, we next used GRSs in pheWAS analyses. In the overall pheWAS analysis with the best-performing GRS, we found an association with cervical cancer codes (C53 and D06, p= p=2.1 × 10^-14^ and p=8.9 × 10^-7^), as expected. However, higher genetic risk was also associated with an increased risk of cervical dysplasia (N87, p=2.6 × 10^-37^), viral warts (B07, p=1.9 × 10^-7^), viral agents as the cause of diseases classified to other chapters (B97, p=2.0 × 10^-12^), but also with diseases with a suspected autoimmune etiology: thyroiditis (E06, p=4.8 × 10^-8^) and psoriasis (L40, p=1.1 × 10^-5^). At the same time, a higher cervical cancer GRS was associated with lower risk of lichen planus (L43, p=6.4 × 10^-8^) and other superficial mycoses (B36, p=1.7 × 10^-7^). In sex-stratified analyses, the female-only results largely reflected those of the overall analysis (Supplementary Table 12), while in the male-only analysis, the GRS was significantly associated with viral warts (B07,p=1.5 × 10^-7^).

When we explored the phenotypic associations with the HLA GRS and nonHLA GRS, we expectedly found that the HLA GRS was associated with diagnoses where HLA plays a role in the etiopathogenesis - viral warts (B07, p=5.7 × 10^-14^), other superficial mycoses (B36, p=1.3 × 10^-11^), viral agents as the cause of diseases classified to other chapters (B97, p=4.0 × 10^-11^), malignant neoplasm of cervix uteri (C53, p=1.1 × 10^-11^), vitamin B12 deficiency anemia (D51, p=6.1 × 10^-6^), thyroiditis (E06, p=3.7 × 10^-7^), psoriasis (L40, p=8.5 × 10^-11^), *lichen planus* (L43, p=2.9 × 10^-9^) and cervical dysplasia (N87, p=3.6 × 10^-29^). nonHLA GRS, on the other hand, was associated only with different cervical phenotypes - cervical dysplasia (N87, p=2.4 × 10^-10^), cervicitis (N72, p=1.5 × 10^- 7^), erosion and ectropion of cervix uteri (N86, p=2.7 × 10^-7^). In the nonHLA GRS pheWAS, cervical cancer codes (C53 and D06) were nominally significant but did not pass the multiple testing threshold (p=2.1 × 10^-4^ and p=1.2 × 10^-4^, respectively).

## Discussion

In this study, we present the results from the largest multi-ancestry GWAS meta-analysis of cervical cancer and other cervical phenotypes, including up to 9,229 cervical cancer cases and 490,304 controls. Compared to the latest cervical cancer GWAS meta-analysis ^8^, our study is larger and has a multi-ancestry approach, includes a wider selection of phenotypes, uses multiple post-GWAS analyses to finemap the signals, and explores the utility of a genetic risk score for cervical cancer. We report four strong non-HLA signals (*LINC00339/CDC42/CDC42-AS1, PAX8/PAX8-AS1, CLPTM1L, GSDMB*), and leverage the latest computational methods and available genomics datasets to pinpoint the most likely causal genes and variants for each associated locus. We further exploit the genetic similarity between cervical cancer and dysplasia to refine the association signal in the *HLA* locus and propose a potential association on chromosome 19 near *CD70*, which can be followed up in further validation studies. By analysing the genetics of cervical ectropion and cervicitis in addition to dysplasia and cancer, we conclude that *PAX8*/*PAX8-AS1* appears to have a dual role in cervical biology: PAX8 signalling is not only important for female genital system development, but *PAX8* is also upregulated in reproductive cancers, enhancing the proliferation of tumor cells ^52^. Previously, it has been reported that several novel *PAX8* transcripts can be observed in cervical carcinoma, indicating differential regulation properties during carcinogenesis ^53^.

Cervical dysplasia is the first step towards cervical malignancy. Overall, the identified genetic associations were very similar in both dysplasia and cancer, and mirrored closely the results from a recent joint analysis of severe dysplasia and cervical cancer ^8^. This indicates that further studies could include both phenotypes to increase power to detect novel associations. One exception was rs12611652 near *DAPL1/PKP4*, which was associated with dysplasia but not with cancer. Given that *DAPL1* has a role in epithelial differentiation, apoptosis, and is potentially a suppressor of cell proliferation, and *PKP4*, highlighted in colocalization analysis, is associated with invasion and metastasis of cancer ^54^, both genes are interesting candidates for further analysis because of their potential protective effect in cervical malignancy development.

Our study provides additional support for potential causal variants and genes at each locus. Although previous studies have reported relevant association signals, they have not mapped the most likely causal genes and variants at each locus, which is an important step in understanding the underlying biology. Evaluating the colocalisation of GWAS signals from different traits (including gene expression) gives valuable information on potential shared causal variants, providing the necessary link between genetics, gene expression and disease risk. We were able to detect colocalisation with gene expression or transcription events for all the evaluated non-HLA loci, which provides evidence that variants in our GWAS signal are involved in regulating the expression or transcription of these genes. At the same time, since reproductive tissues are underrepresented in widely used gene expression datasets, we had to rely on tissues that are similar to female reproductive tract tissues, based on cellular composition and gene expression patterns. Therefore, more extensive characterisation of gene expression regulation in reproductive tract tissues is urgently needed to facilitate correct interpretation of GWAS signals. We also constructed the 95% credible sets of causal variants and compared them to chromatin annotations in HeLa cervical carcinoma cell line to evaluate which of these could potentially be most relevant for cervical cancer. Our analysis shows that several credible set variants overlap either with transcription start sites or enhancer regions, providing additional support for potential causal variants. Considering recent studies, it is plausible that there are several tightly linked causal variants in each locus ^55^ and functional studies are needed for further fine-mapping.

Overall, our results are in line with previous findings by replicating the associations near *PAX8* ^7, 8^, *CLPTM1L* ^8^, *HLA-DRB1* ^46^, *HLA-B* ^8^, and *GSDMB* ^42^. The association on chromosome 1 appears to be novel in the context of cervical pathology, although the region is a known risk locus for other gynecological problems, such as endometriosis, uterine fibroids, pelvic organ prolapse and ovarian cancer. Our results support *LINC00339* and *CDC42/CDC42-AS1* as the most likely candidate genes in this locus, which is in line with evidence from other cancers ^33–36^. In fact, previous studies have shown that knocking down *LINC00339* expression leads to increased *CDC42* expression ^56^, which is supported by data from eQTLs – variants associated with increased expression of *LINC00339* have an opposite effect on *CDC42* expression ^57^. Thus, it cannot be ruled out that several jointly regulated genes in this locus contribute to cancer pathogenesis. The value of our study is best highlighted by the *GSDMB* locus, which has been associated with cervical cancer previously, but the signal has not been dissected in detail, thus the underlying mechanisms have remained unclear. We show the signal includes a splice acceptor variant that has a direct impact on *GSDMB* functionality - the rs11078928-C allele prevents the splicing of exon 6 and consequently suppresses the pyroptotic activity of the GSDMB protein. GSDMB is involved in antitumour immunity and tumour suppression ^58^, meaning that suppression of GSDMB activation would lead to increased tumor progression, which is in line with the results of our study, where the C-allele is associated with an increased risk of cervical cancer. At the same time, there is evidence that the role of GSDMs in cancer might be context-specific and thus more studies are needed to evaluate the role of GSDMB in cervical epithelium immunity and cervical cancer, and potential therapeutic perspectives ^58^.

The GRS constructed based on our analyses shows strong association with cervical cancer in the EstBB. Additional analyses show that a large part of the predictive power comes from associations in the HLA region, which is not surprising given the major role of HPV infection and HLA-mediated immune response in the pathogenesis of cervical malignancy. Although further analyses are needed, this might indicate that in the context of cervical cancer, testing of HLA alleles might be largely sufficient for risk profiling. On the other hand, this also underlines the importance of considering disease biology when constructing GRS and using appropriate LD reference panels, since many commonly used LD references offer different coverage for the HLA region ^59^ and may not capture the correct population-specific LD structure, therefore leading to underperformance of tested GRS. A previous study exploring the performance of GRS in cervical cancer found that women in the highest 5% have approximately 22% risk of developing cervical neoplasia ^60^; however, in this study the cases and controls originated from different populations, which can lead to unwanted stratification and differences in allele frequencies, making it difficult to compare with our results. More recent studies ^7, 61^ constructed a GRS for cervical cancer using ten variants (all on chr6) with corresponding ORs from previous literature and validated its association with cervical cancer with an OR=1.22 per standard deviation increase in the GRS, which is similar to what we observe for the HLA fraction of our GRS in the discovery set, and HR=1.22, which is less than what we see for our risk score (HR=1.33) in the validation set. Collectively, these results indicate that a GRS for cervical cancer captures the genetic risk well and might be useful for research and screening purposes in the future. A recent paper ^62^ highlighted main avenues how population-level screening could be improved by including GRS. First, GRSs may improve the identification of individuals who would benefit from inclusion in screening programs, or the timing of screening initiation (in the context of cervical cancer screening, this may mean that for women with high genetic risk, screening might start earlier or continue for an extended period). Second, women with high genetic risk might benefit from more frequent screening, and third, GRS might be useful in selecting the tools (in the context of cervical cancer, either HPV testing or PAP smear analysis) used as part of screening. To inform these decisions, more detailed analyses on the relationship between cervical cancer GRS and HPV infection and/or pathology histological features are needed.

A pheWAS with the GRS demonstrated a positive association with other diagnoses associated with HPV infection (cervical dysplasia and viral warts) and HLA involvement (psoriasis, thyroiditis), and a negative association with *lichen planus* and superficial mycoses. The etiology of *lichen planus* is somewhat poorly studied and potentially involves autoimmune etiology, but a decreased incidence of cervical cancer has been demonstrated in *lichen sclerosus* (another skin disease with suspected autoimmune etiology and preference for the genitalia ^63^) patients ^64^. Although the genetics of *lichen planus* has not been studied thoroughly, our results suggest that in terms of HLA associations, cervical malignancy and lichen planus are mirror phenotypes.

It has been suggested that persistent HPV infection and cervical cancer are more common in women with autoimmune disease ^65, 66^, partly because of systemic immunosupressive drugs prescribed to these women ^65^. However, our results suggest shared genetic predisposition may also play a role, as the combination of HLA alleles associated with risk of cervical dysplasia has also been associated with autoimmune diseases. This is supported by the association we see between the cervical cancer GRS and autoimmune conditions (psoriasis, thyroiditis). Together, these results further support targeted HPV vaccination in women with autoimmune conditions ^67^.

The conducted genetic correlation analyses indicating significant genetic correlation between cervical cancer and age at first birth and smoking closely mirror the results from a recent Mendelian randomization analysis, which showed smoking increases, and older age at first pregnancy decreases the risk of cervical cancer, respectively ^8^. Several potentially interesting correlations (such as with parental age at death and lung cancer) were nominally significant in our analysis and future studies with larger sample sizes and more power are needed to fully elucidate the shared genetics between these traits and cervical malignancy. At the same time, the method (LDSC) commonly used for genetic correlation analysis on summary level data excludes the HLA region which is a major contributor to cervical cancer heritability, therefore other complementing approaches (such as exploring HLA allele pleiotropy) should be employed to fully understand the genetic correlations between cervical cancer and other traits.

Our analyses are based on population-based biobank data, which offers access to large sample sizes, but at the same time it can hinder the accessibility to more detailed clinical information (such as HPV status), especially when using summary-level data. Further studies evaluating the detected loci in relation to specific HPV strains or histopathological features will elucidate their more specific role in cervical pathology etiopathogenesis. We used relatively simple phenotype definitions based solely on ICD-codes, which on one hand simplifies data analysis, but on the other hand may introduce unwanted heterogeneity as the use of these codes might somewhat vary in different healthcare systems. However, we replicate many previously reported associations with cervical cancer, suggesting our approach is suitable. Although our study is the first attempt at a multi-ancestry GWAS meta-analysis, demonstrating similar effect estimates in both analysed ancestries, the number of non-European samples is small, and given the high prevalence of cervical malignancy in non-European populations, additional Black and Asian populations should be included in analyses to also improve the transferability of genetic risk scores.

Our study provides the most comprehensive genetic risk assessment of different cervical phenotypes to date. We provided the first insight into the genetics of cervical ectropion and cervicitis, which is an important step towards a complete understanding of cervical biology. We further clarify the genetic background of cervical malignancy, supporting the involvement of genes important for reproductive tract development, immune response, and cellular proliferation/apoptosis. The detailed characterisation of association signals, together with mapping of causal variants and genes, and the construction of a GRS offers valuable leads for further functional studies which may eventually lead to better treatment and prevention of cervical neoplasia.

## Data Availability

FinnGen R5 data can be browsed and downloaded from https://r5.finngen.fi. Biobank Japan data can be browsed and downloaded from http://jenger.riken.jp:8080. GWAS meta-analysis summary statistics will be made available upon publication of this manuscript.

http://jenger.riken.jp:8080

## Acknowledgements

This study was supported by European Union through Horizon 2020 grant INTERVENE, the Estonian Research Council grants PRG687, PRG1291 and PSG776 and by MATER Marie Sklodowska-Curie which received funding from the European Union’s Horizon 2020 research and innovation program under grant agreement No. 813707. Computational analyses of Estonian Biobank data were performed in the High Performance Computing Center, University of Tartu. U.V was funded by the European Regional Development Fund and the programme Mobilitas Pluss (MOBTP108).

The FinnGen project is funded by two grants from Business Finland (HUS 4685/31/2016 and UH 4386/31/2016) and the following industry partners: AbbVie Inc., AstraZeneca UK Ltd, Biogen MA Inc., Bristol Myers Squibb (and Celgene Corporation & Celgene International II Sàrl), Genentech Inc., Merck Sharp & Dohme Corp, Pfizer Inc., GlaxoSmithKline Intellectual Property Development Ltd., Sanofi US Services Inc., Maze Therapeutics Inc., Janssen Biotech Inc, and Novartis AG. Following biobanks are acknowledged for delivering biobank samples to FinnGen: Auria Biobank (www.auria.fi/biopankki), THL Biobank (www.thl.fi/biobank), Helsinki Biobank (www.helsinginbiopankki.fi), Biobank Borealis of Northern Finland (https://www.ppshp.fi/Tutkimus-ja-opetus/Biopankki/Pages/Biobank-Borealis-briefly-in-English.aspx), Finnish Clinical Biobank Tampere (www.tays.fi/en-US/Research_and_development/Finnish_Clinical_Biobank_Tampere), Biobank of Eastern Finland (www.ita-suomenbiopankki.fi/en), Central Finland Biobank (www.ksshp.fi/fi-FI/Potilaalle/Biopankki), Finnish Red Cross Blood Service Biobank (www.veripalvelu.fi/verenluovutus/biopankkitoiminta) and Terveystalo Biobank (www.terveystalo.com/fi/Yritystietoa/Terveystalo-Biopankki/Biopankki/). All Finnish Biobanks are members of BBMRI.fi infrastructure (www.bbmri.fi). Finnish Biobank Cooperative -FINBB (https://finbb.fi/) is the coordinator of BBMRI-ERIC operations in Finland. The Finnish biobank data can be accessed through the Fingenious^®^ services (https://site.fingenious.fi/en/) managed by FINBB.

## Author contributions

Data analysis: Mariann Koel, Urmo Võsa, Maarja Lepamets, Kristi Läll, Triin Laisk Writing: Mariann Koel, Urmo Võsa, Natàlia Pujol-Gualdo, Priit Palta, Reedik Mägi, Triin Laisk

Data: Estonian Biobank Research Team, FinnGen, Susanna Lemmelä, Hannele Laivuori, Mark Daly

Group authorship: Estonian Biobank Research Team, FinnGen

## Supplementary Tables

**Supplementary Table 1.** Number of cases and controls for each analysed phenotype

| Dataset | Ectropion | Cervicitis | Dysplasia | Cervical cancer |
| --- | --- | --- | --- | --- |
| EstBB | 9,664/82,378 | 18,192/73,850 | 10,448/81,594 | 748/81,869 |
| FinnGen R5 | 498/68,969 | 1,093/111,858 | 4,246/68,969 | 1,313/99,048 (R4) |
| Rashkin et al | NA | NA | NA | 5,998/189,855 (UKBB)<br>565/29,801 (Kaiser) |
| Biobank Japan | NA | NA | NA | 605/89,731 |
| **Total** | 10,162/151,347 | 19,285/185,708 | 14,694/150,563 | 9,229/490,304 |

**Supplementary Table 2.**
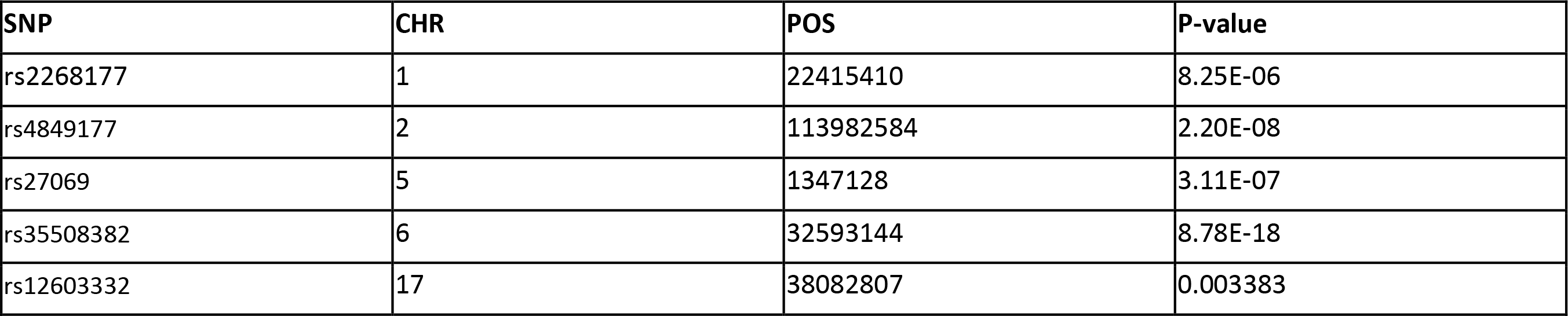
Cervical cancer GWAS meta-analysis lead signals in cervical dysplasia GWAS meta-analysis

| SNP | CHR | POS | P-value |
| --- | --- | --- | --- |
| rs2268177 | 1 | 22415410 | 8.25E-06 |
| rs4849177 | 2 | 113982584 | 2.20E-08 |
| rs27069 | 5 | 1347128 | 3.11E-07 |
| rs35508382 | 6 | 32593144 | 8.78E-18 |
| rs12603332 | 17 | 38082807 | 0.003383 |

## Supplementary Figures

**Supplementary Figure 1.**
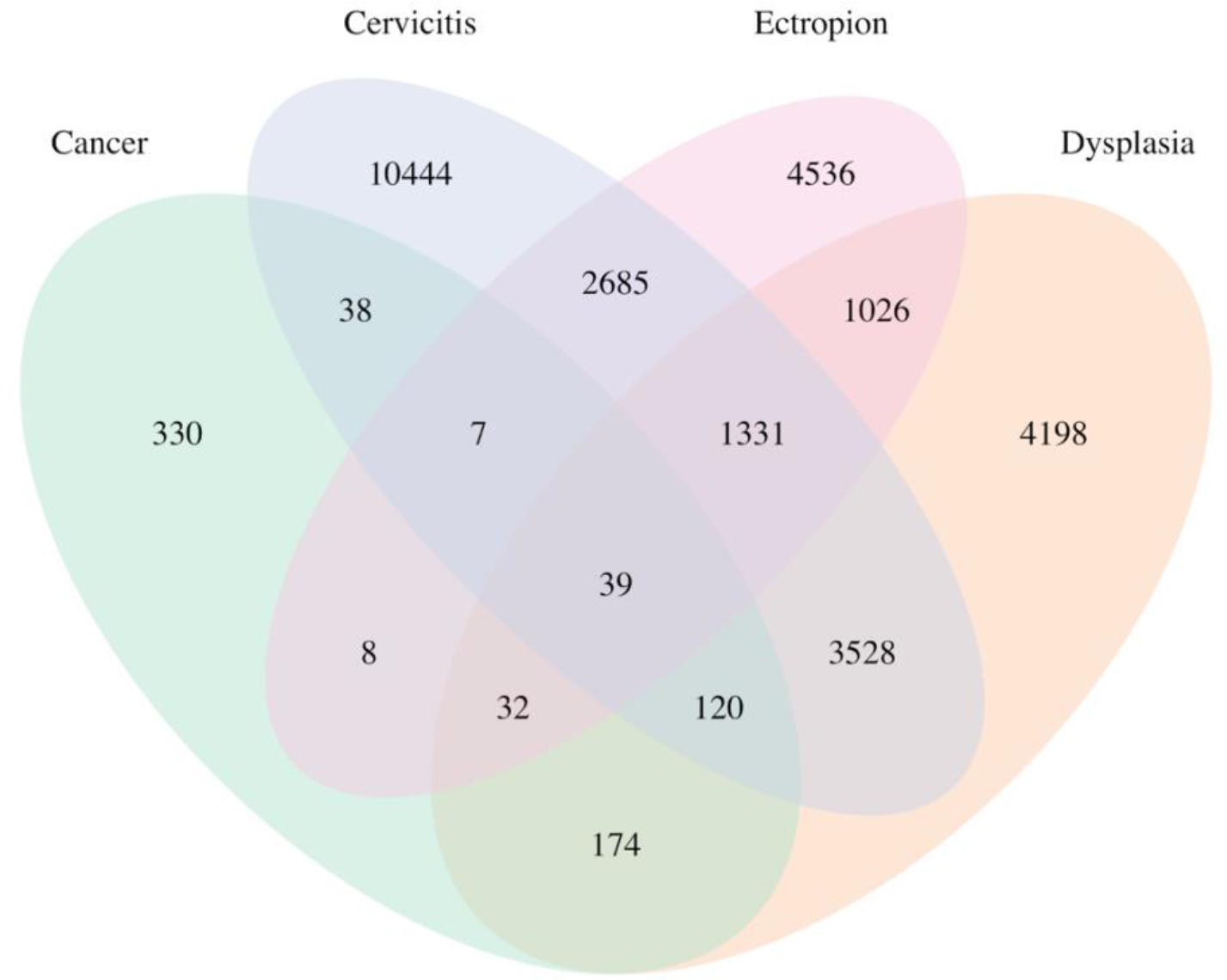
Venn diagramm showing overlap in different diagnoses related to uterine cervix in the Estonian Biobank.

**Supplementary Figure 2.**
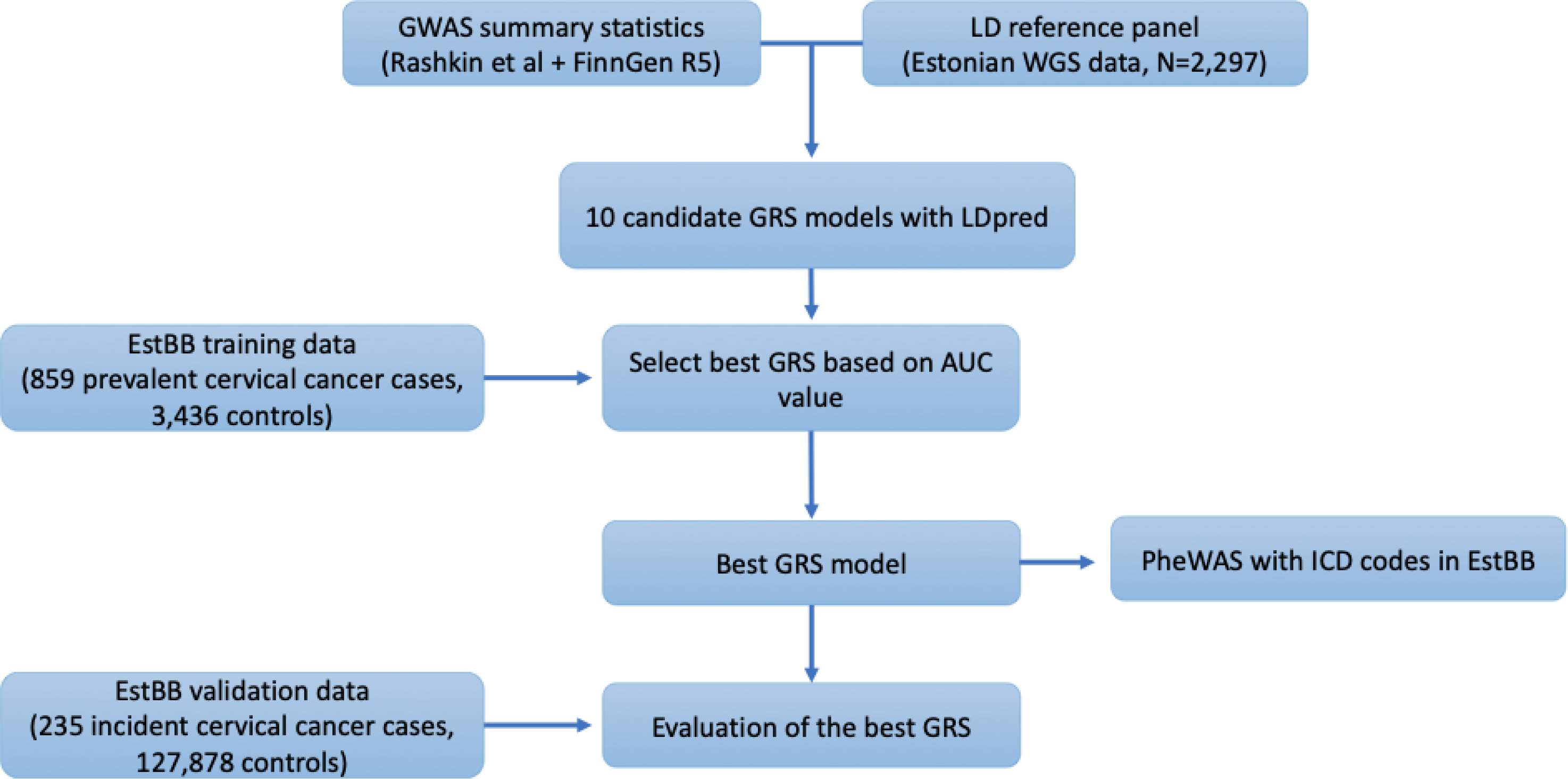
Workflow for GRS calculation and validation in EstBB.

**Supplementary Figure 3.**
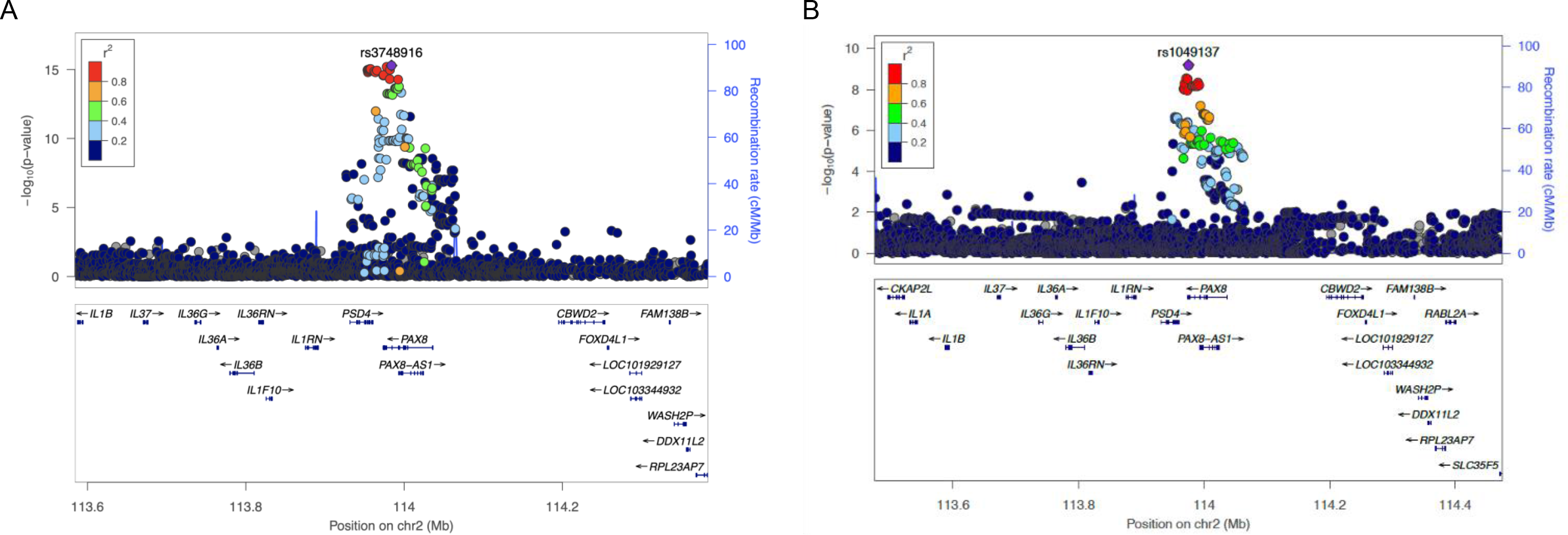
Regional plots for cervical ectropion (A) and cervicitis (B) GWAS meta-analysis signals on chromosome 2.

**Supplementary Figure 4.**
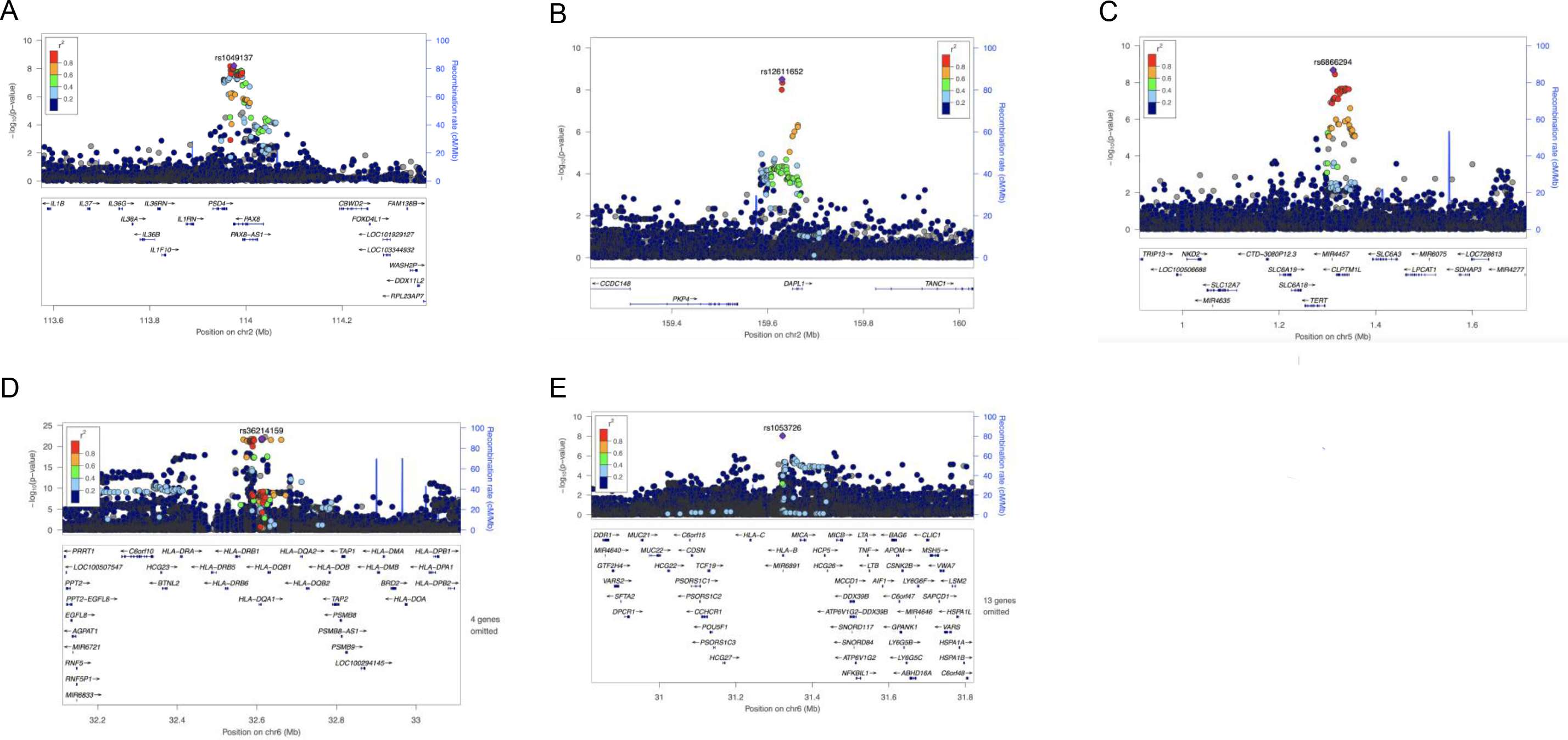
Regional plots for cervical dysplasia GWAS meta-analysis signals on chr2 (A, B), chr5 (C) and chr6 (D, E)

**Supplementary Figure 5.**
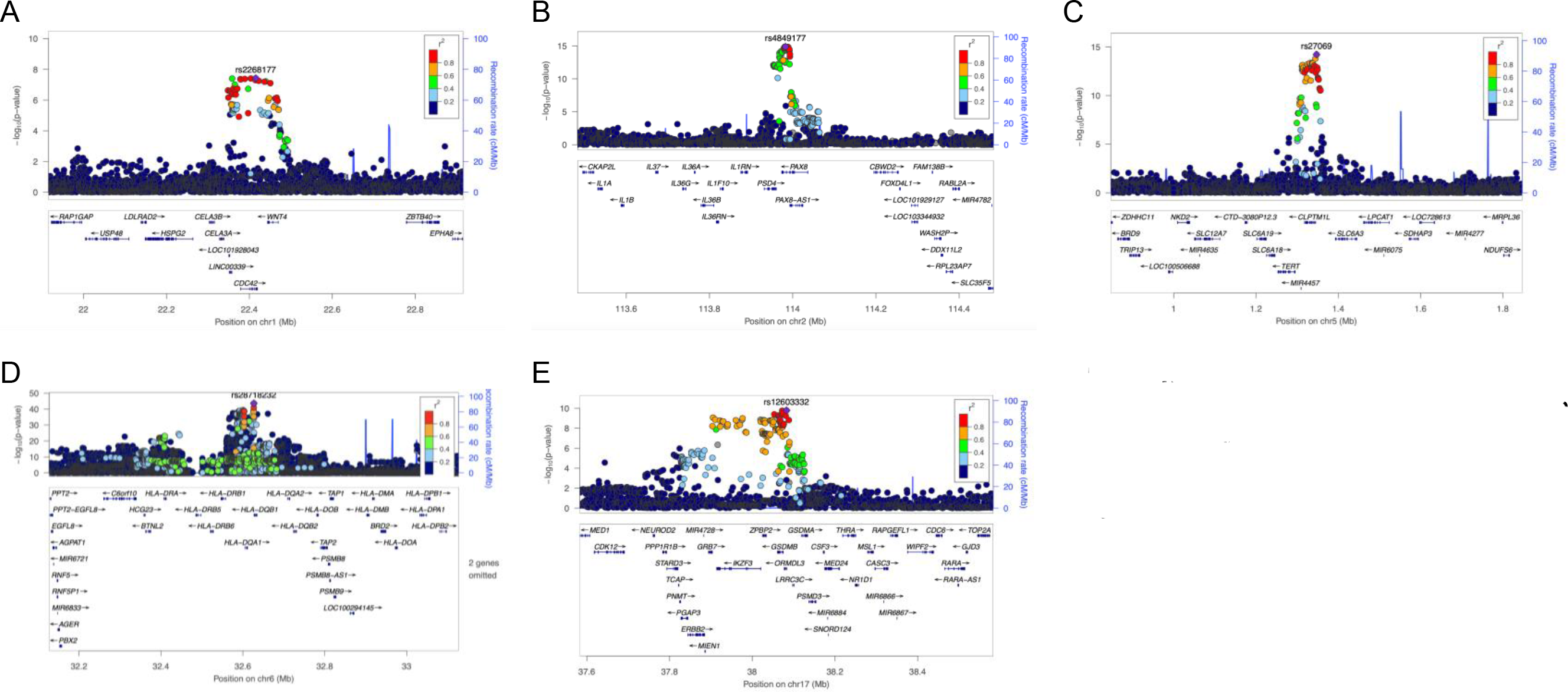
Regional plots for cervical cancer European ancestry GWAS meta-analysis signals on chr1 (A), chr2 (B), chr5 (C), chr6 (D) and chr17 (E).

**Supplementary Figure 6.**
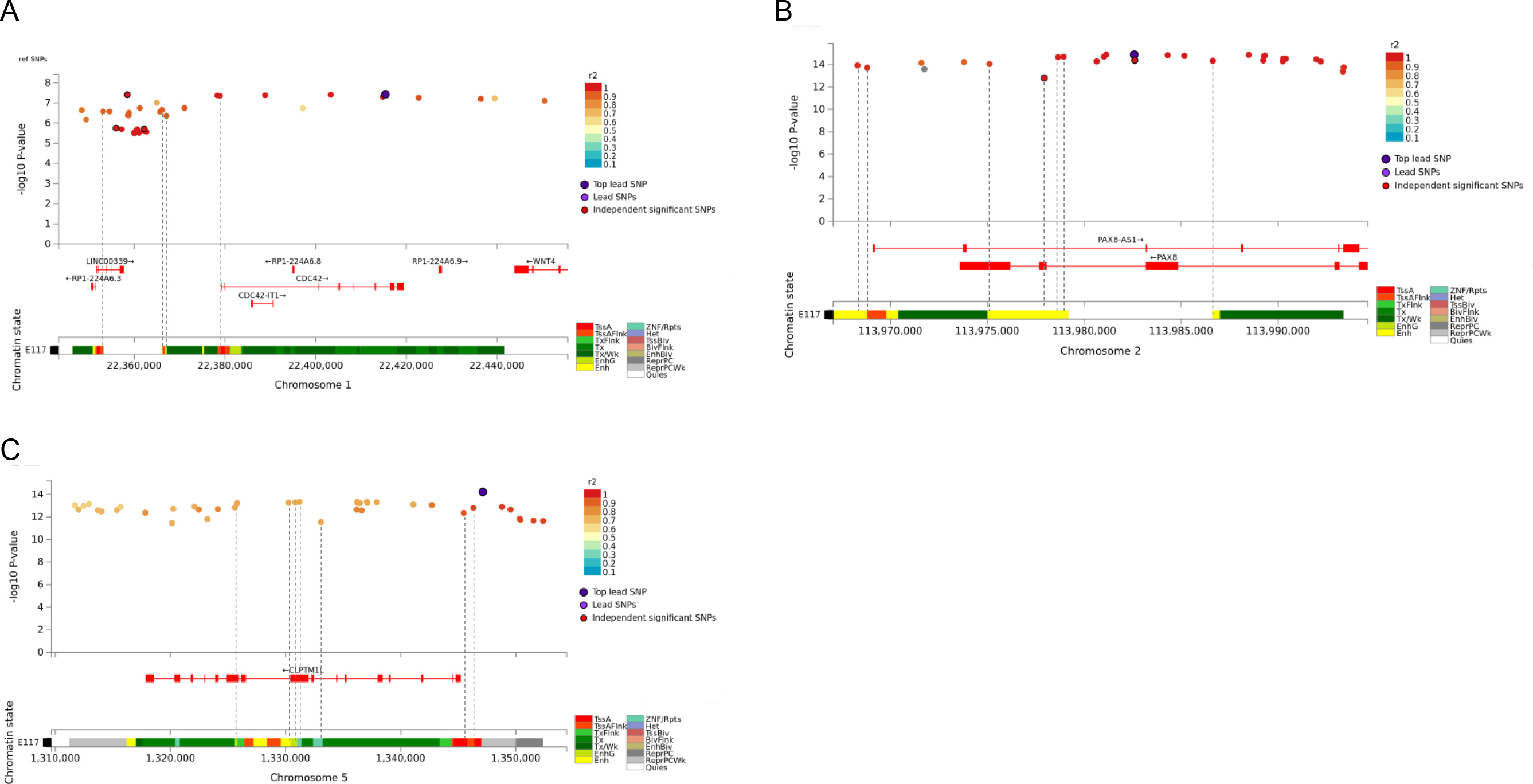
Cervical cancer GWAS meta-analysis credible sets for chr1 (A), chr2 (B), and chr5 (C), and their overlap with chromatin annotations in HeLa cell line (Encode Roadmap)

**Supplementary Figure 7.**
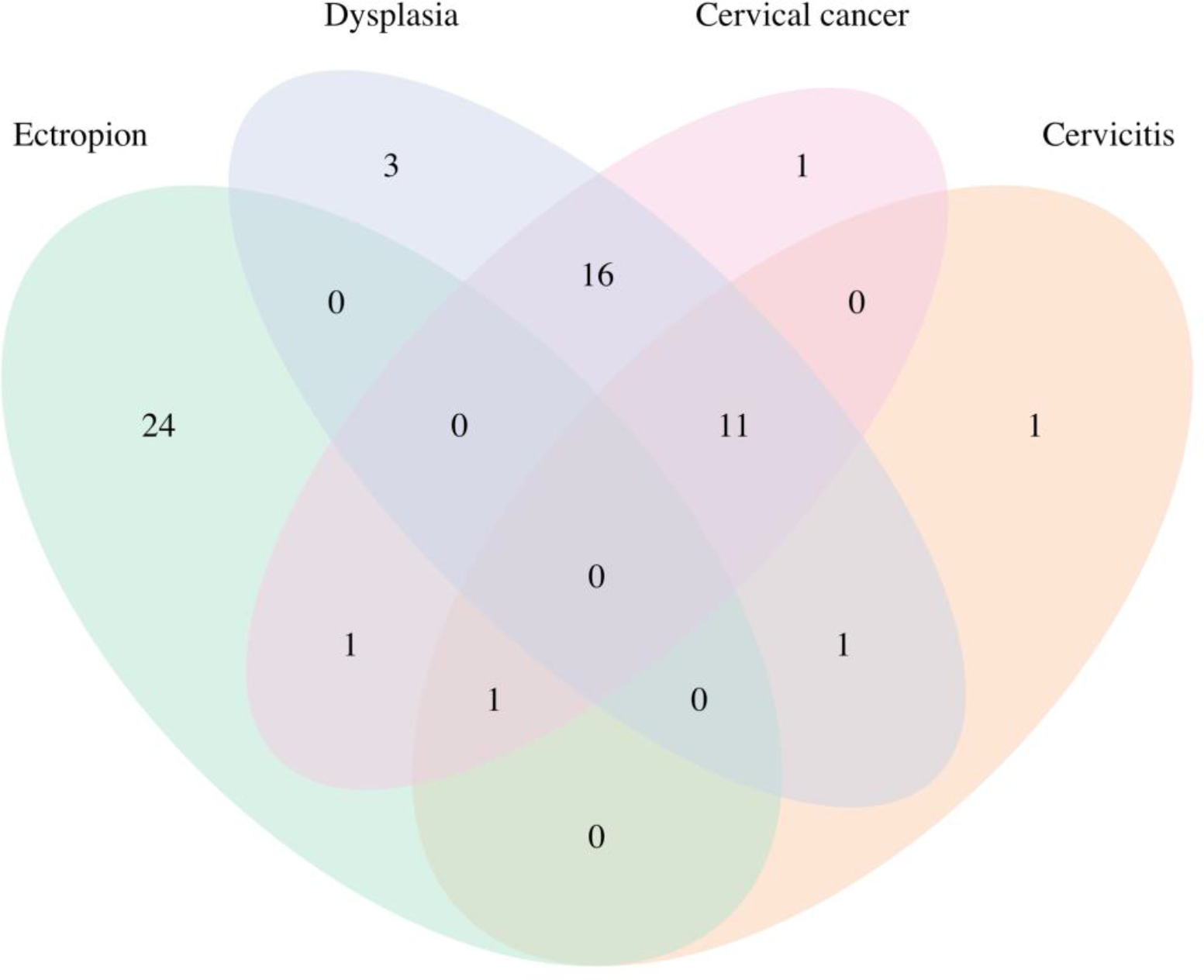
Venn diagram illustrating the overlap between credible set SNPs in cervical ectropion, cervicitis, dysplasia and cervical cancer

**Supplementary Figure 8.**
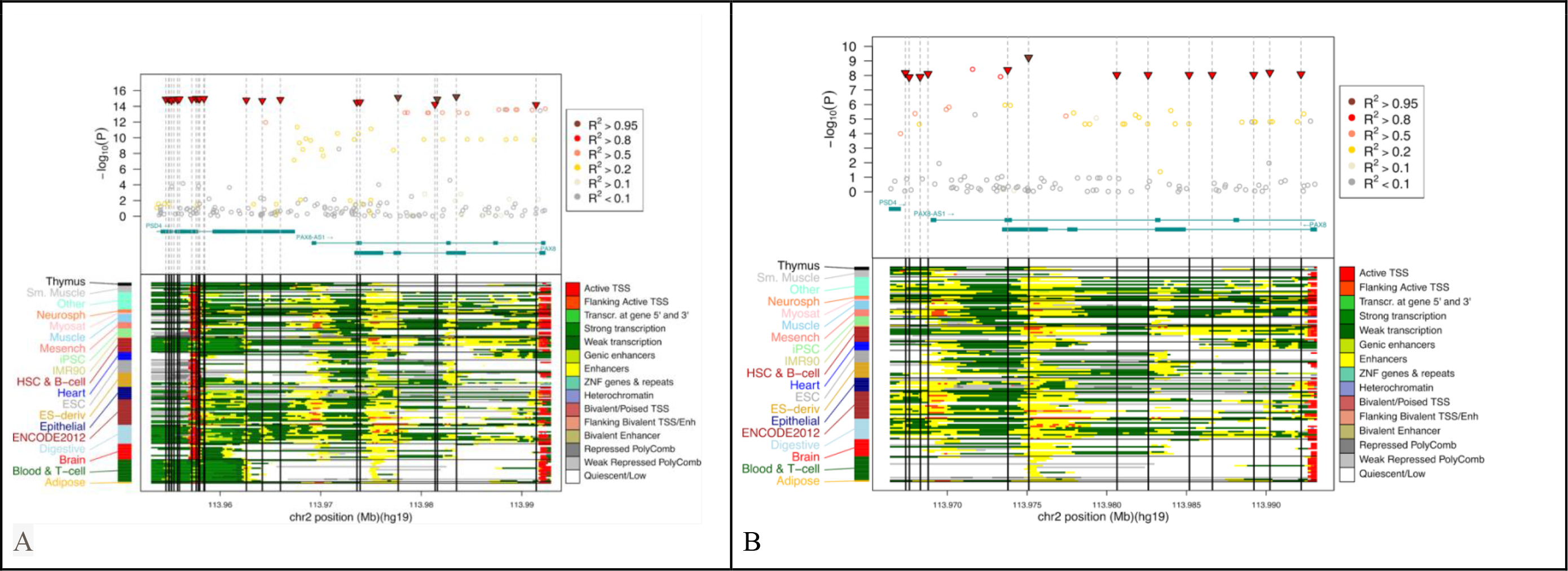
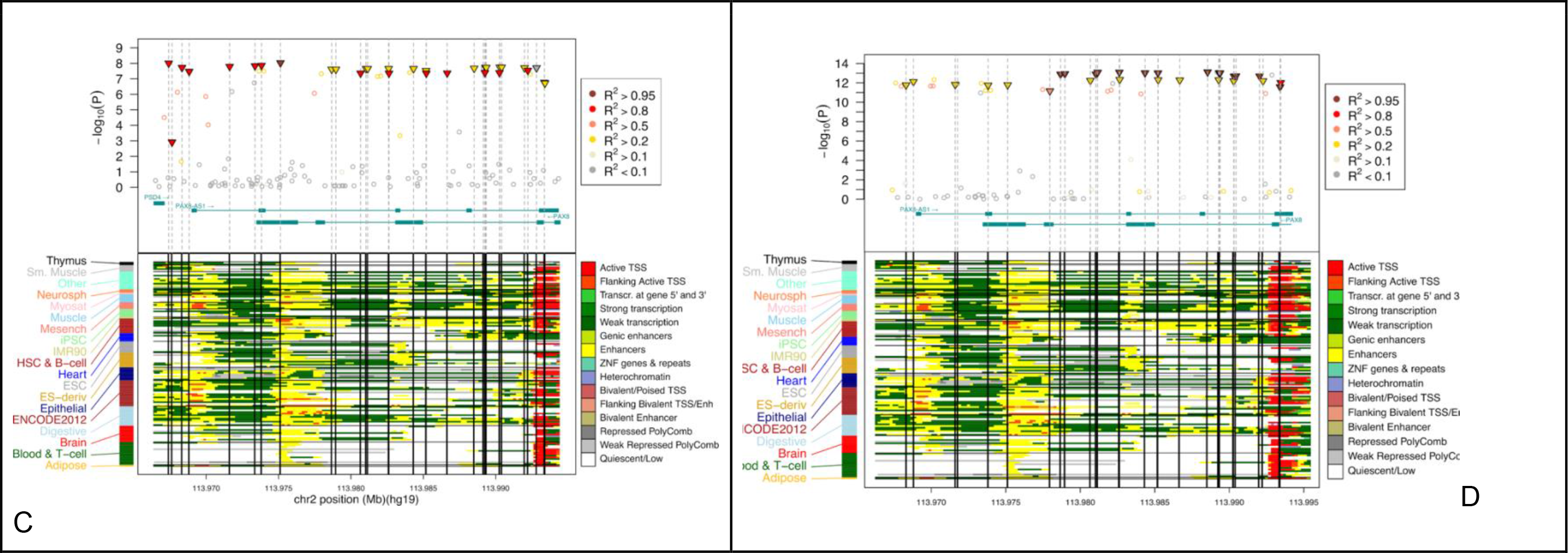
Regional plots highlighting HyPrColoc credible set SNPs for chr2 locus in cervical ectropion (A), cervicitis (B), dysplasia (C) and cervical cancer (D) GWAS analysis. Bottom panel shows the corresponding Roadmap Epigenomics chromatin annotations in different tissues.

**Supplementary Figure 9.**
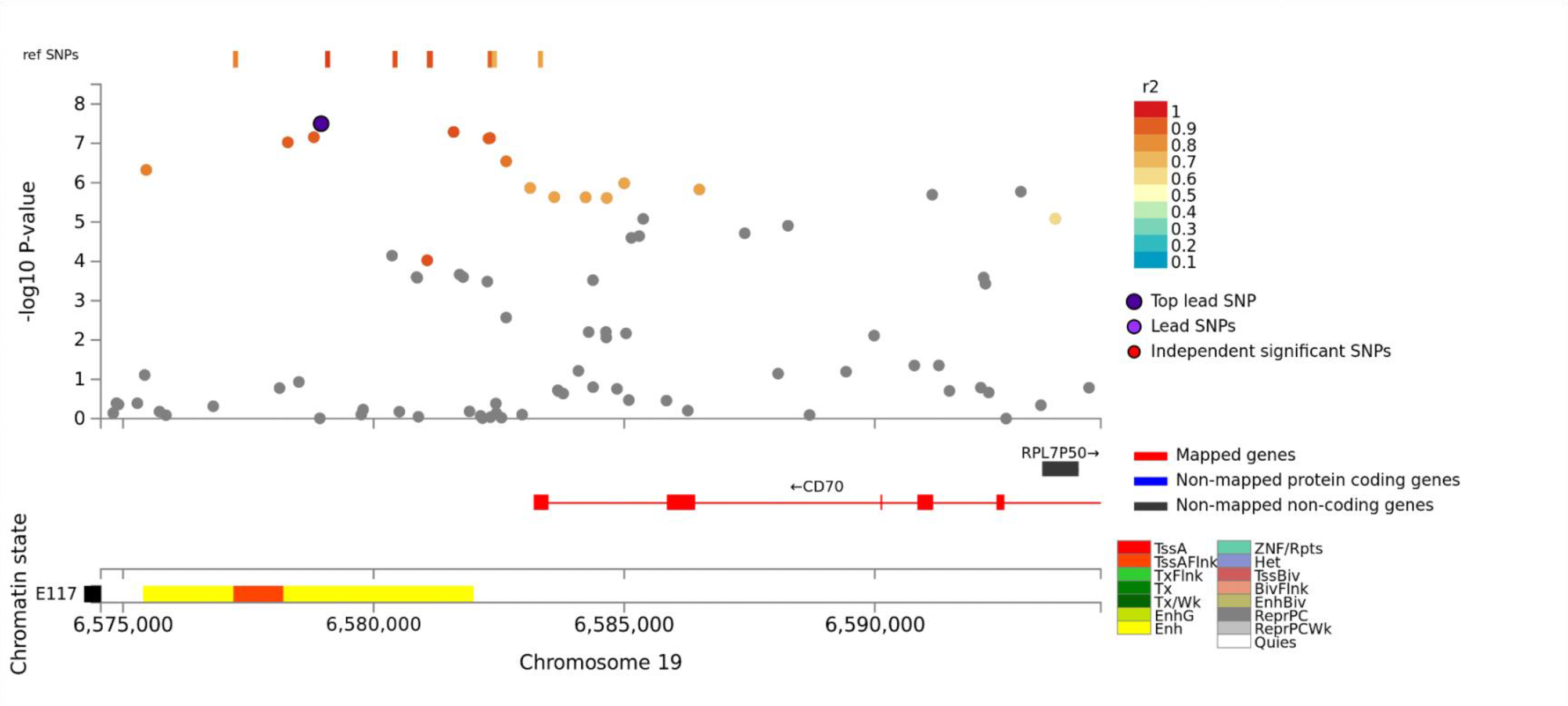
Regional plot for locus on chromosome 19 identified in MTAG joint analysis of cervical dysplasia and cancer.

**Supplementary Figure 10.**
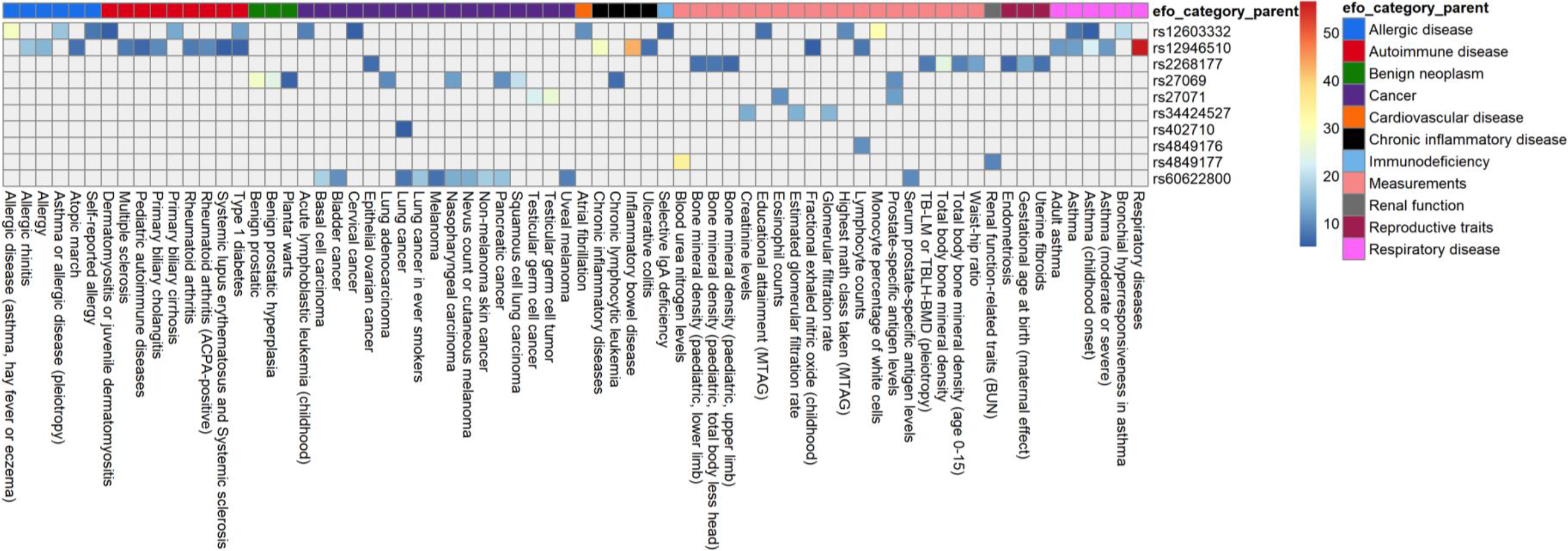
Results of the GWAS catalog look-up for cervical cancer loci

**Supplementary Figure 11.**
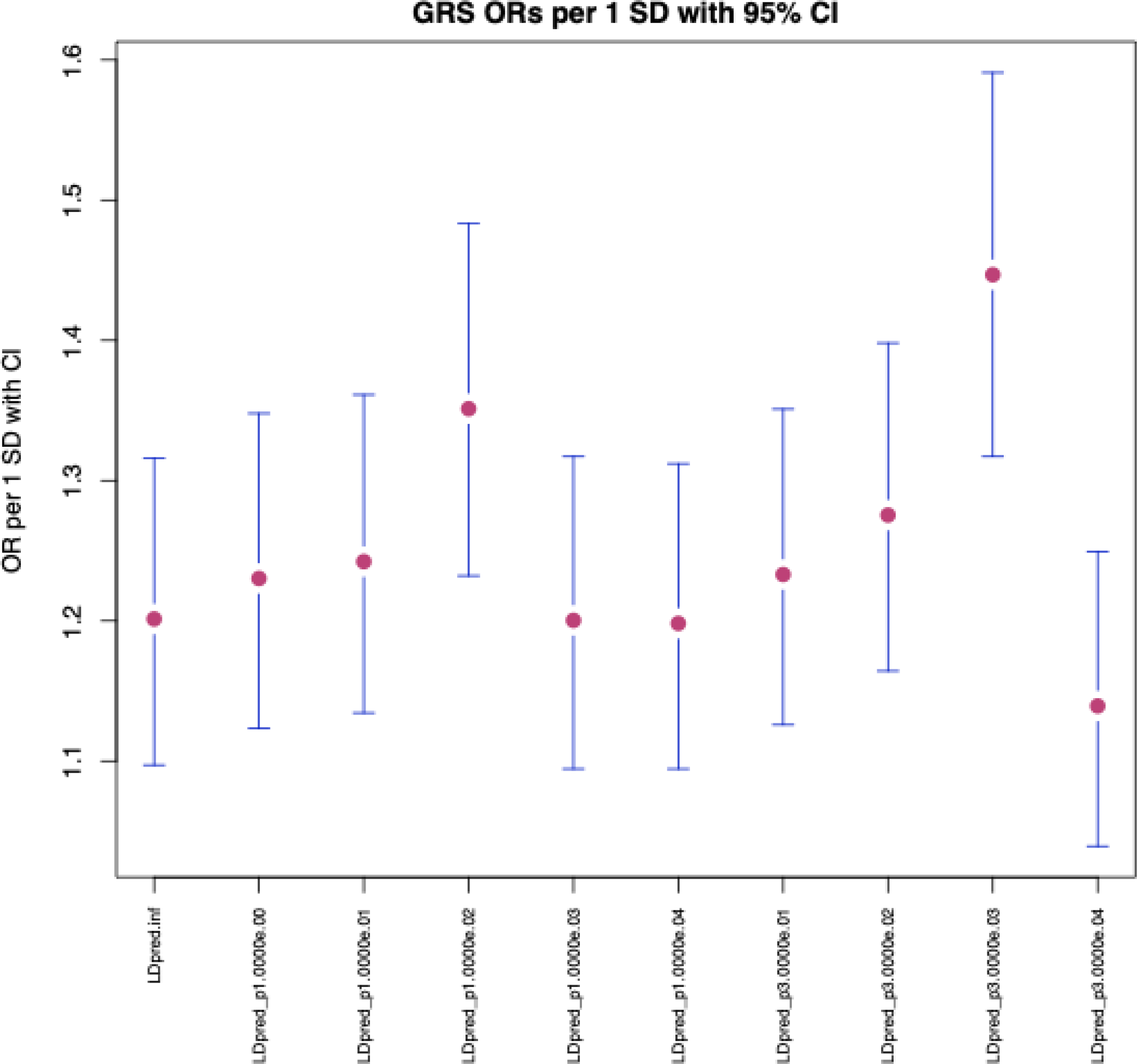
Odds ratios (dots) and 95% CI-s (error bars) for association with cervical cancer diagnosis in discovery set for different cervical cancer genetic risk scores (x-axis).

